# Development and evaluation of a method for classifying foods to be consumed in moderation

**DOI:** 10.1101/2025.03.20.25324317

**Authors:** Amara Channell Doig, Leah M. Lipsky, Allison Choe, Tonja R. Nansel

## Abstract

**Background:** While the 2020-2025 Dietary Guidelines for Americans (DGA) recommend limiting foods high in added sugars, saturated fat, refined grains, and sodium, they do not provide criteria for classifying foods high in these components.

**Objective:** To develop and evaluate quantitative criteria for classifying moderation foods informed by the DGA.

**Design:** The 2017-2018 Food and Nutrient Database for Dietary Studies (FNDDS, n=6909 foods) was used to evaluate food-level content validity, by examining the proportion of moderation foods within recommended and non-recommended food groups, and convergent validity, by comparing nutrient density in moderation versus non-moderation foods. Dietary recall data from the cross-sectional, nationally representative National Health and Nutrition Examination Survey (2017-2018) were used to assess diet-level convergent validity, by investigating correlations of moderation food intake (% kcal) with overall diet quality.

**Participants/Setting:** Participants in the U.S. aged 2+ years (n=6180) were included.

**Main outcome measures:** The Nutrient Rich Foods 9.3 index (NRF) measured food-level nutrient density; Healthy Eating Index-2020 scores (HEI) measured person-level diet quality.

**Statistical Analyses:** T-tests and ANOVA evaluated differences in nutrient density by moderation food classification. Pearson’s correlations estimated associations of moderation food intake (% kcal) with HEI.

**Results:** More non-recommended (e.g., 97% of snacks, 96% of sweets) than recommended (e.g., 19% of vegetables, 23% of fruits) food groups were classified as moderation foods. Mean nutrient density was significantly lower in moderation foods than non-moderation foods (mean difference in NRF scores = −74.8, p<0.001). Moderation food intake (% kcal) was negatively correlated with HEI (r=-0.72), HEI-moderation (r=-0.56), and HEI-adequacy (r=-0.67).

**Conclusions:** The moderation food classification method demonstrated strong content and convergent validity, and may facilitate diet quality assessment, surveillance, and public health interventions.

## Introduction

The 2020 – 2025 Dietary Guidelines for Americans (DGA) recommend limiting intake of foods and beverages high in added sugars, saturated fat, refined grains, and sodium, along with limiting alcoholic beverages^1^ due to associations with adverse health outcomes. ^2–12^ These foods are often energy-dense and may displace intake of nutrient-rich foods needed to meet nutritional requirements (“core foods”). ^1^ However, while the DGA specify recommended overall intake limits for these dietary components, they do not provide criteria for identifying foods to limit.

Given that people select and consume foods, not nutrients, establishing evidence-based criteria for classifying foods that are high in dietary components to limit would facilitate accessible, food-based dietary guidance and nutrition surveillance, improve research on diet and health outcomes, and inform intervention targets to improve diet quality. ^13^

Existing a priori metrics for assessing diet or food quality, such as diet quality indices and nutrient profiling methods, do not provide a standardized food-level approach to identify foods high in DGA dietary components to limit. For example, the Healthy Eating Index 2020 (HEI), a measure of alignment of overall diet quality with the DGA, includes four moderation components for refined grains, sodium, added sugar, and saturated fat, which are scored at the overall intake level (per 1,000 kcal or as a percentage of total energy). ^14^ However, the HEI lacks food-level classification criteria for identifying foods that should be consumed in moderation, i.e., “moderation foods.” Similarly, nutrient profiling methods, such as the Nutrient Rich Food Index (NRF) 9.3^15^ and the Food Compass score, ^16,17^ calculate nutrient-based scores for the purpose of ranking individual foods by overall nutritional value, but these have been based on heterogeneous nutrient criteria, reference amounts, and weighting schemes, and not all methods designate cut-offs for classifying foods.

While several approaches for classifying foods by dietary components to limit have been developed for alternative purposes, several aspects limit their applicability for classifying foods in U.S. food and nutrition databases. For example, food classification approaches developed to assist charitable food organizations in procuring healthy foods,^18–21^ may involve complicated and inconsistent criteria, incomplete food lists, or manual inspection of individual food labels or ingredient lists. U.S. regulatory and policy frameworks, including the U.S. Food and Drug Administration (FDA) “healthy” or “high in” labeling criteria, ^22,23^ address most dietary components to limit (excluding refined grains and alcohol), but were developed to regulate food manufacturers’ labeling practices and do not apply to the full range of food groups, non-packaged foods, or mixed foods consumed by the U.S. population. Furthermore, FDA thresholds are based on the FDA-defined serving size and 20% daily values for a fixed 2,000-kcal reference intake level, rather than food-specific nutritional profiles, ^22,23^ which may obscure nutrient density and pose challenges for informing a consistent method to identify foods high in dietary components to limit. Additionally, FDA thresholds are based on the FDA-defined serving size and 20% daily values for a fixed 2,000-kcal reference intake level, rather than food-specific nutritional profiles, ^22,23^ which may obscure nutrient density and pose challenges for informing a consistent method to identify foods high in dietary components to limit. For example, according to the FDA definition, plain sugar would not be considered high in added sugar since a serving size of 1 tsp contains 16 kcal, which is less than 20% of the daily value for a 2,000-kcal diet. While several countries have also developed front-of-package labeling thresholds, these are informed by country-specific dietary guidance and priorities and are not aligned with DGA-based recommendations. ^24,25^

A standardized, evidence-based approach for identifying moderation foods that is applicable to food- and dietary intake databases could enable consistent application across studies and facilitate direct translation of dietary guidance into actionable intervention targets and public health initiatives to improve diet quality in the US. Therefore, this study aimed to (1) develop quantitative criteria for classifying moderation foods informed by the DGA that can be applied to existing food and dietary intake databases for research, and (2) evaluate its ability to differentiate foods according to nutrient density and distinguish between diets of high versus low quality.

## Methods

This work proceeded in two phases. First, development of the Moderation Food Classification Method (MFCM) involved selecting, testing, and refining criteria for each dietary component used to classify foods as either moderation or non-moderation. Following selection of the final criteria, the MFCM was evaluated at both the food and diet levels to examine its content, construct, and convergent validity (**Figure 1**). Content validity was assessed as the extent to which the MFCM aligned with the concept of moderation foods as described in the peer-reviewed literature. ^26^ Construct validity was evaluated by examining the extent to which the MFCM was consistent with its underlying theoretical framework, specifically whether the classifications accurately captured dietary components to limit. ^27^ Convergent validity was assessed as the extent to which the MFCM aligned with other measures with theoretically related measures, including nutrient density and diet quality. ^26^

**Figure 1.**
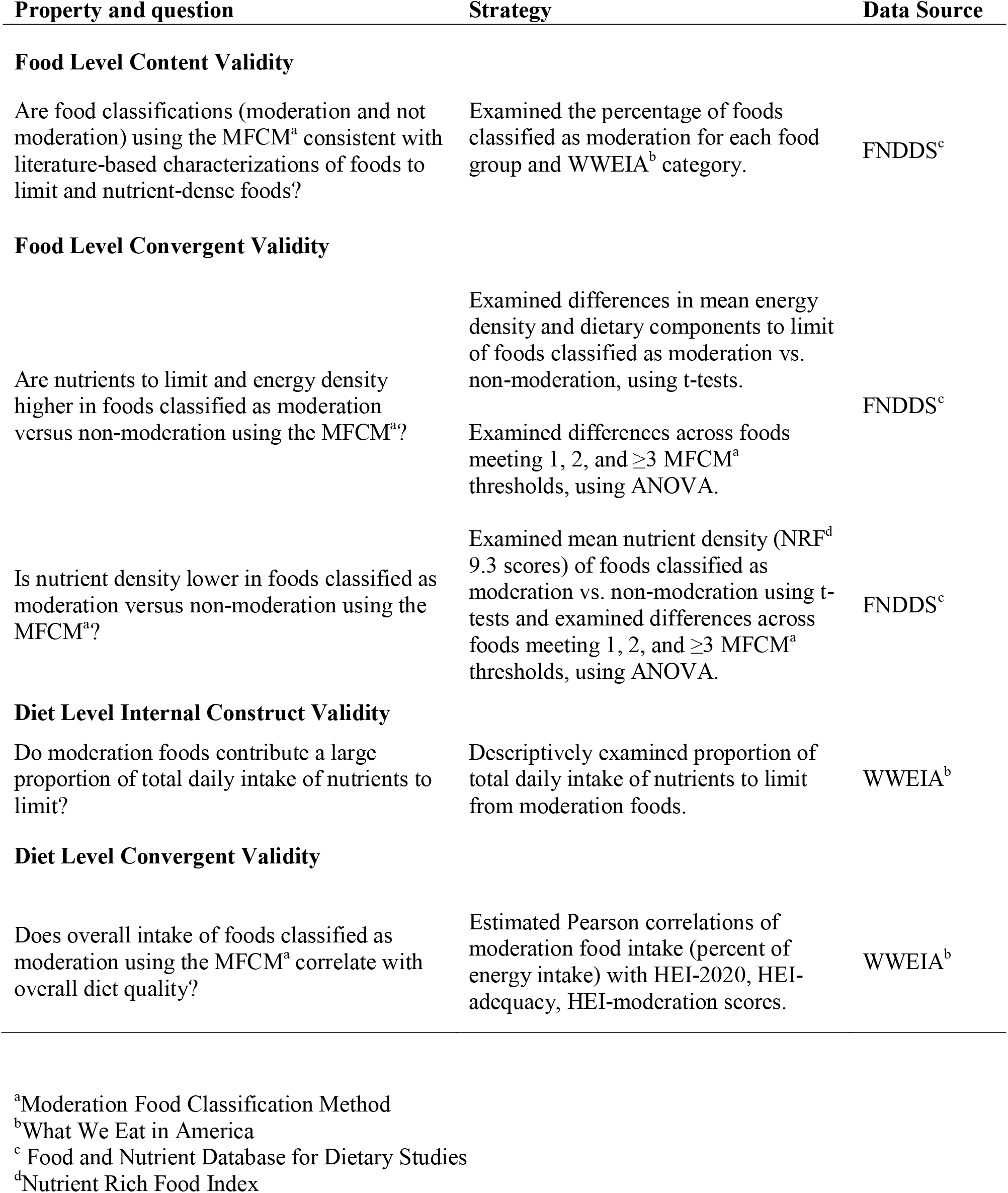
Approaches used to evaluate the Moderation Food Classification Method.

### Process for developing the Moderation Food Classification Method (MFCM)

Three guiding principles were used for developing the MFCM, including (1) alignment with the dietary component recommendations identified in the DGA, ^1^ (2) alignment with food group categorizations in the DGA and peer-reviewed literature, (3) parsimony and applicability to the Food and Nutrient Database for Dietary Studies (FNDDS) ^28^ food codes and dietary intake data collected in the What We Eat in America (WWEIA) National Health and Examination Survey (NHANES) dietary recalls^29^ or using the National Cancer Institute Automated Self-Administered 24-hour dietary recall application (ASA24). ^30^ The first principle was operationalized by aiming to use DGA-based criteria to identify foods containing high amounts of dietary components to limit in the DGA^1^ and referred to as moderation components in the HEI^14^ (i.e., added sugars, saturated fat, sodium, and refined grains). The second principle was operationalized by aiming to classify as moderation foods those that are consistently described in the scientific literature as moderation, discretionary, junk, or extra (e.g., sweets, salty snacks, sugar sweetened beverages, high sugar cereal, processed meats, fried potatoes, and high-fat fast food, as outlined in **Figure 2**), ^31–45^ while not classifying as moderation those foods described as nutrient-dense (e.g., “vegetables, fruits, whole grains, seafood, eggs, beans, peas, and lentils, unsalted nuts and seeds, fat-free and low-fat dairy products, and lean meats and poultry—when prepared with no or little added sugars, saturated fat, and sodium”). ^1^ The third principle was operationalized by aiming to develop criteria that can be calculated from variables currently available in the FNDDS and ASA24 data and applied across all food groups within these data sources, deviating from established criteria or creating food-group specific criteria only if required to ensure consistency with the first two principles.

**Figure 2.**
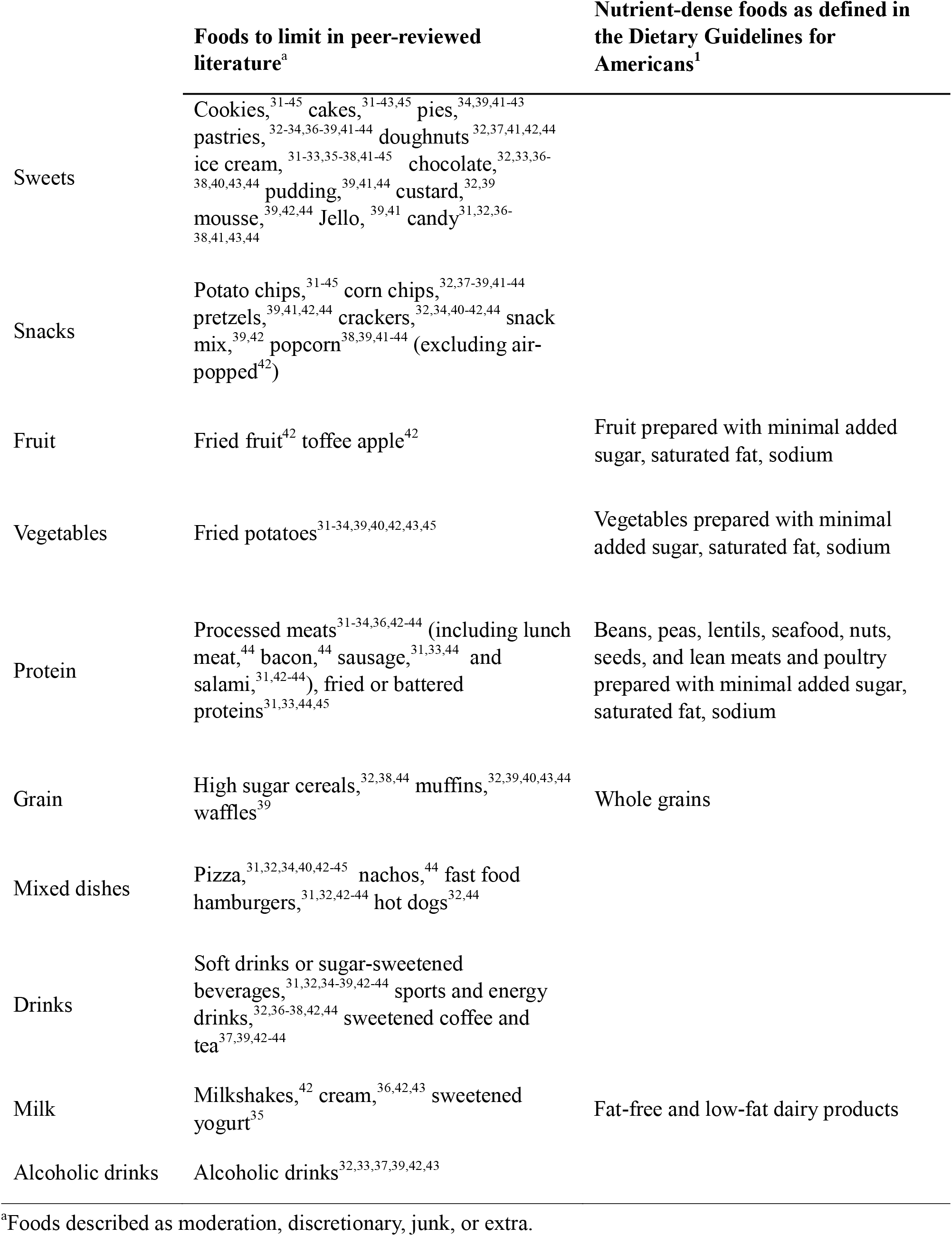
Foods defined as foods to limit or nutrient dense for measurement development and content validity.

### Criteria development

Aligned with the first principle, initial criteria were selected in accordance with values of dietary components to limit referred to in the DGA^1^ and the HEI^14^ scoring algorithm. In alignment with the second principle, initial criteria were evaluated with respect to their accuracy in classifying foods to limit and nutrient-dense foods within each food group. Since the HEI^14^ and the DGA^1^ were developed for evaluating overall dietary intake, the initial criteria based on these guidelines resulted in some misclassifications at the individual food level. The initial criteria were then modified accordingly, by incorporating food-group specific criteria or additional established criteria, where appropriate, in accordance with the third principle. Each of the criteria tested and a summary of the findings are presented in **Figure 3**.

**Figure 3.**
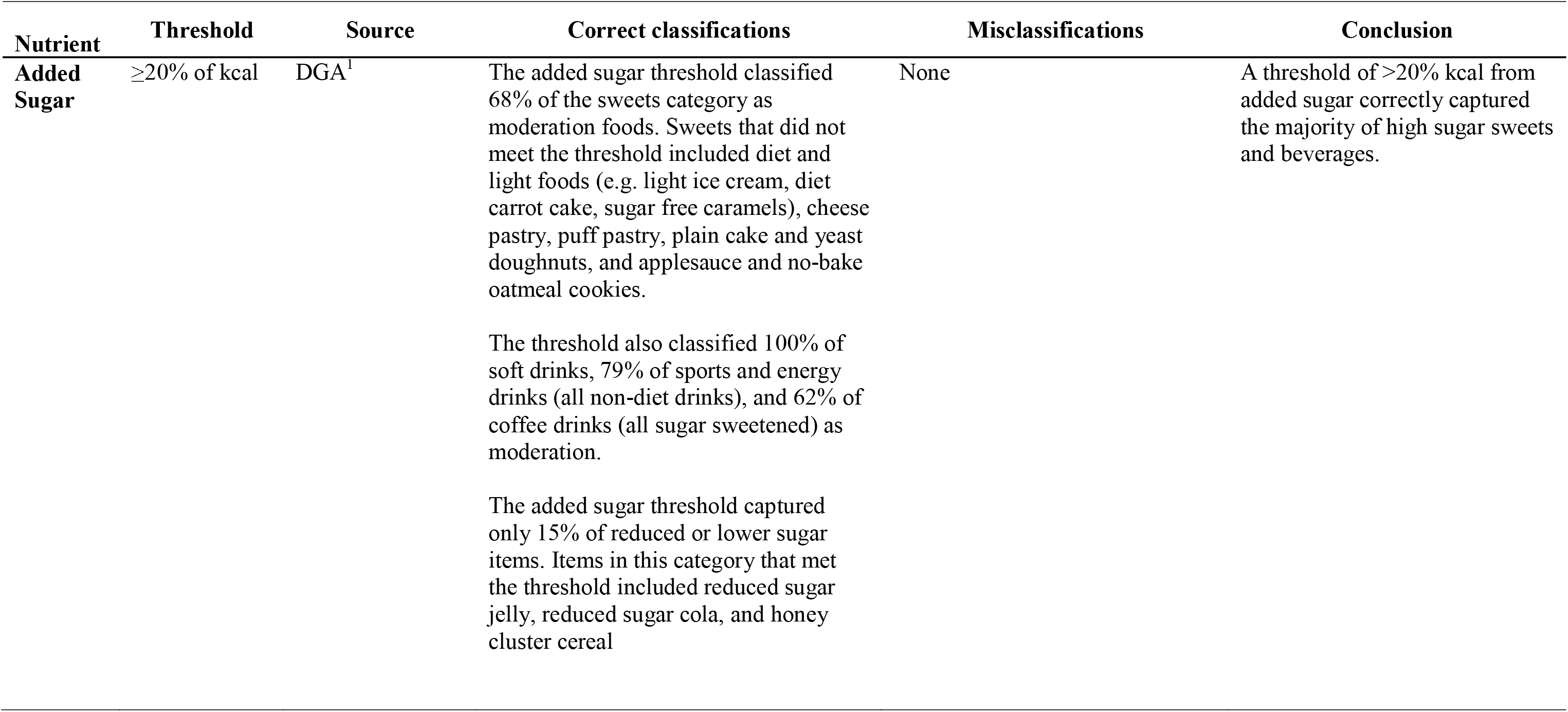
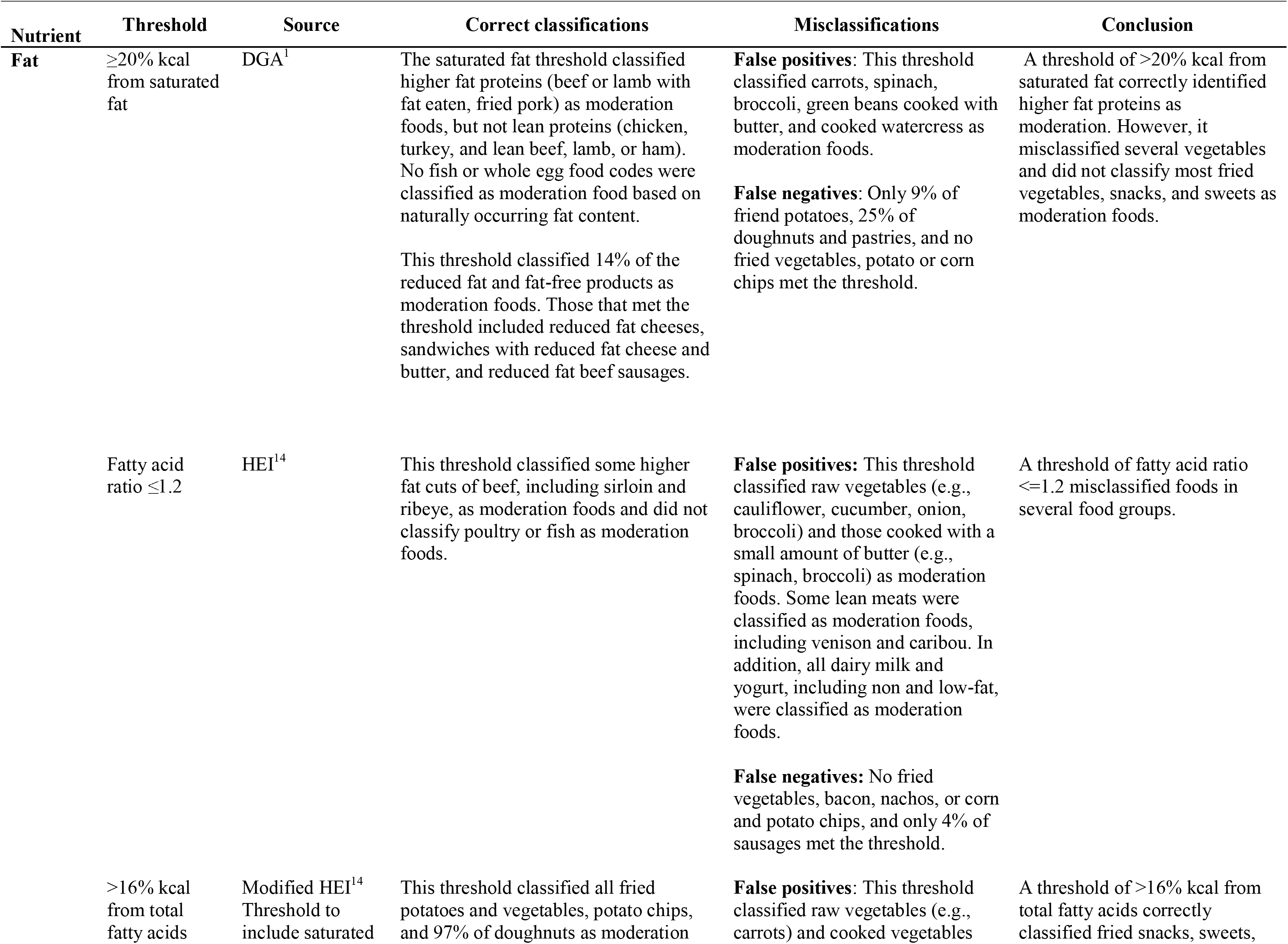
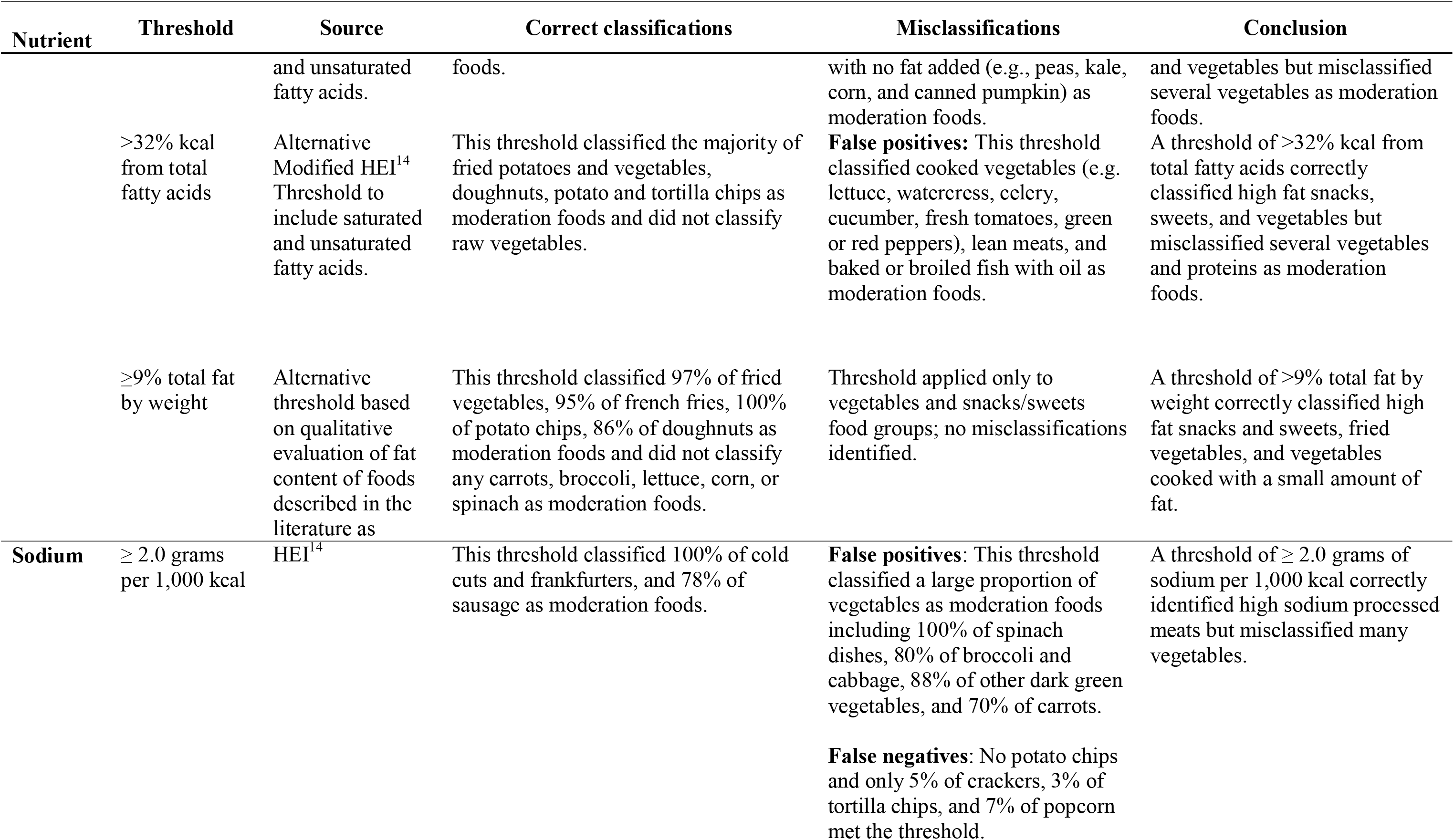
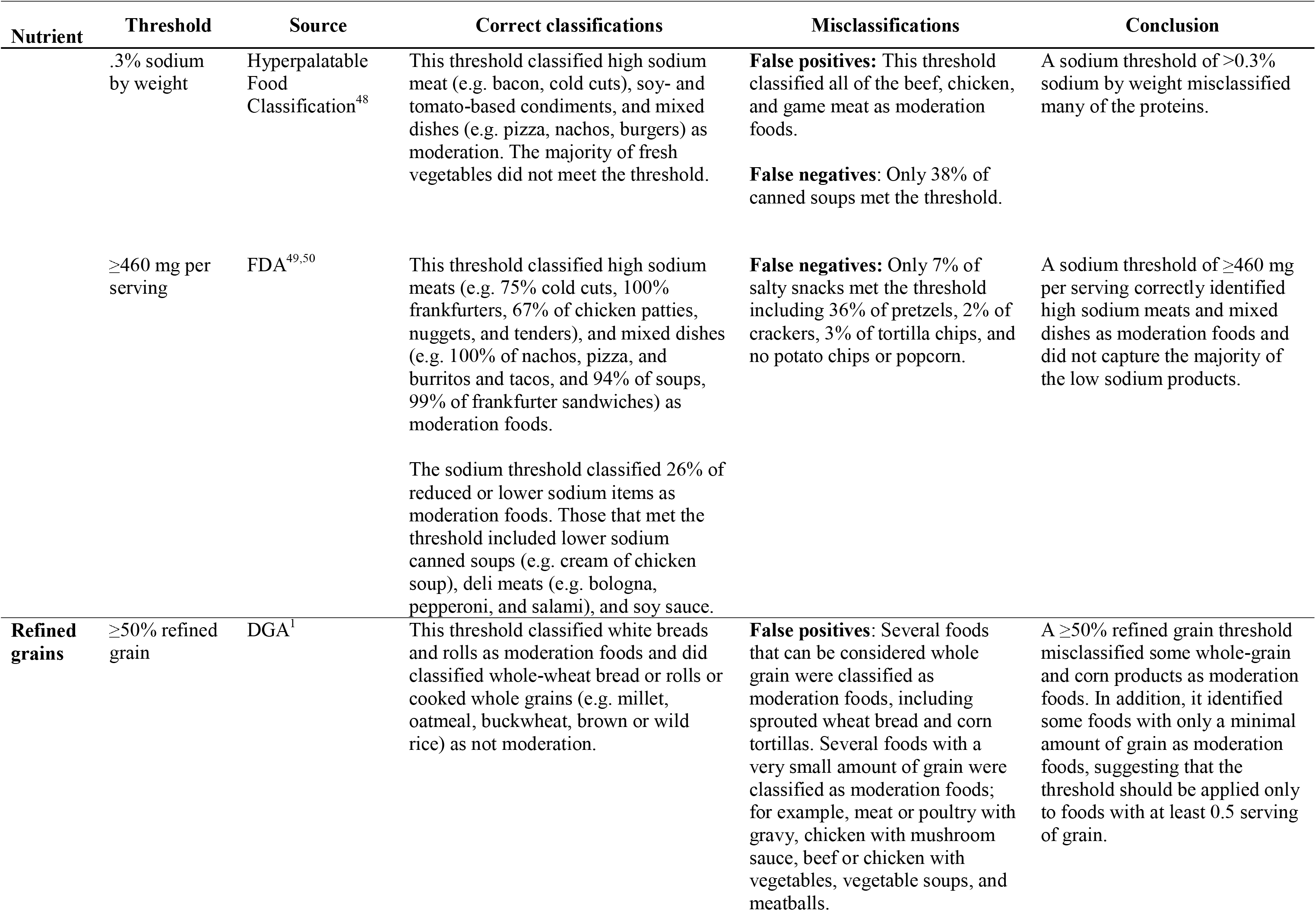
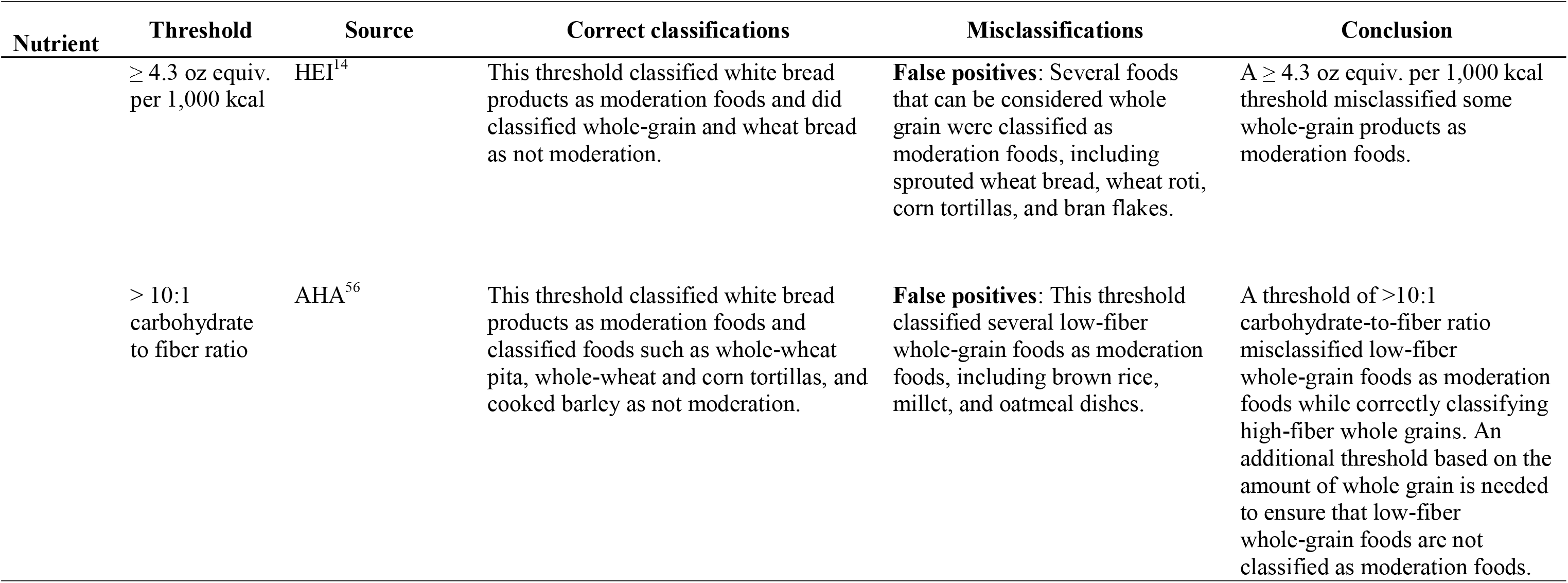
Moderation Food Classification Method criteria tested, classification findings, and decisions.

#### Added Sugars

The DGA recommends that added sugars contribute less than 10% of total daily energy intake. ^1^ However, because this recommendation applies to the overall diet rather than to individual foods, foods containing modestly more than 10% of kcal from added sugars may still fit within a healthful dietary pattern. Therefore, the initial criterion for classifying an individual food as high in added sugars was set at ≥20% of kcal from added sugars, or twice the recommended daily dietary-level limit. This criterion correctly classified the majority of the high sugar sweets and sugar sweetened beverages and resulted in no false positives based on DGA-defined nutrient-dense foods (**Figure 2)**, so was maintained.

#### Fats

Using the same rationale as for added sugars, the initial criterion for saturated fat was set to ≥20% of kcal, which is twice the DGA maximum recommended daily intake of saturated fat for the overall daily diet. ^1^ Evaluation of this criterion revealed misclassifications in the vegetables group and the snacks and sweets group based on the second guiding principle. Among vegetables, foods prepared with small amounts of butter (e.g. broccoli, collards, or carrots cooked with butter) were classified as moderation foods (false positives) because their inherently low energy density caused even small amounts of added saturated fat to exceed the criterion.

Conversely, many fried vegetables (e.g., fries, potato tots, onion rings, vegetable fritters), snacks (e.g., potato chips, corn chips), and sweets (e.g., doughnuts) were not classified as moderation foods (false negatives) because fat added during preparation was primarily unsaturated (oils) and, thus, not captured by the saturated fat criterion. Together, these misclassifications indicate that the saturated fat criterion did not appropriately characterize foods containing added fat in these food groups. Although current dietary guidelines do not limit total or unsaturated fat intake, they recommend consuming foods in nutrient-dense forms and identify less nutrient-dense foods as those prepared with additions such as butter, creamy sauces, and frying oil. ^1^ Additionally, the DGA scientific report identifies fried vegetables and chips as foods associated with lower diet quality and higher adiposity, ^46^ and these foods are consistently referred to in the literature as discretionary, junk, extra, or moderation. ^31–45^ A criterion based on the quantity of fat added during preparation (e.g., in frying) would have been optimal, but this is not feasible using variables within the FNDDS or FPED. Therefore, total fat was explored as a proxy indicator of added fat within these two food groups (vegetables, snacks and sweets). This approach is also broadly consistent with the USDA Smart Snack Standards, which include a limit for total fat among the nutrient criteria for snacks sold in schools. ^47^ The total fat criterion was restricted to these food groups where misclassification by the saturated fat criterion was high and to minimize the potential for foods naturally high in unsaturated fat to be misclassified as moderation foods (e.g., nuts and seeds). We evaluated multiple alternative approaches to incorporate both saturated and unsaturated fat for the vegetable and snacks/sweets food groups (**Figure 3**). The final criteria that resulted in the strongest alignment with the guiding principles included: ≥ 20% kcal from saturated fat for all food groups other than vegetables, ≥9% total fat by weight for vegetables, and either criterion for snacks/sweets.

#### Sodium

An initial criterion of ≥2.0 grams of sodium per 1,000 kcal was evaluated, based on the value corresponding to the minimum score for the HEI sodium component. ^14^ However, because sodium has no energy value, using an energy-density-based cut point developed for the whole diet resulted in misclassifications (false positives) when applied to individual foods. For example, several vegetables cooked with a small amount of salt exceeded this threshold due to their low energy content (e.g. raw spinach, fresh kale, carrots, green beans, or broccoli cooked with no added fat). After examining multiple alternative criteria using energy or weight as the referent value, the criterion was revised to align with the FDA definition of a high-sodium food (460 mg per serving), which corresponds to 20% of the recommended maximum daily intake for a 2,000-kcal diet (2300 mg). ^49,50^

#### Refined Grains

The initial criterion of ≥50% of total grains from refined grains (oz equivalents), based on the DGA recommendation that half of grains consumed should be whole grains, ^1^ was selected for classifying foods as moderation. However, this criterion resulted in misclassification of sprouted grain bread, corn tortillas, and other foods made from masa as moderation foods (false positives). While these foods are labeled as refined grains in the USDA Food Patterns Equivalents Database (FPED), ^51^ this contradicts guidance from the Special Supplemental Nutrition Program for Women, Infants, and Children, ^52^ the National School Lunch Program, ^53^ and the Child and Adult Care Food Program^54^, which refers to these foods as whole grains. To address this, the final refined grains criteria incorporated the American Heart Association whole grain criterion^55^ (carbohydrate to fiber ratio of <10:1) in addition to the initial DGA-based criterion of ≥50% total grains from refined grains. To avoid misclassifying mixed dishes containing only a small amount of refined grains (e.g., soup thickened with cornstarch), the grain criteria were applied only to foods containing at least 0.5 ounce-equivalents of total grains.

#### Alcohol

Based on the DGA guidance to limit alcohol intake, ^1^ all alcoholic beverages were classified as moderation.

#### Final Criteria

The final criteria for classifying moderation foods are presented in **Figure 4**. Any food meeting at least one component criterion is classified as a moderation food. In addition to the dichotomous moderation food classification system, an ordinal classification value was created by summing the number of component criteria met.

**Figure 4.**
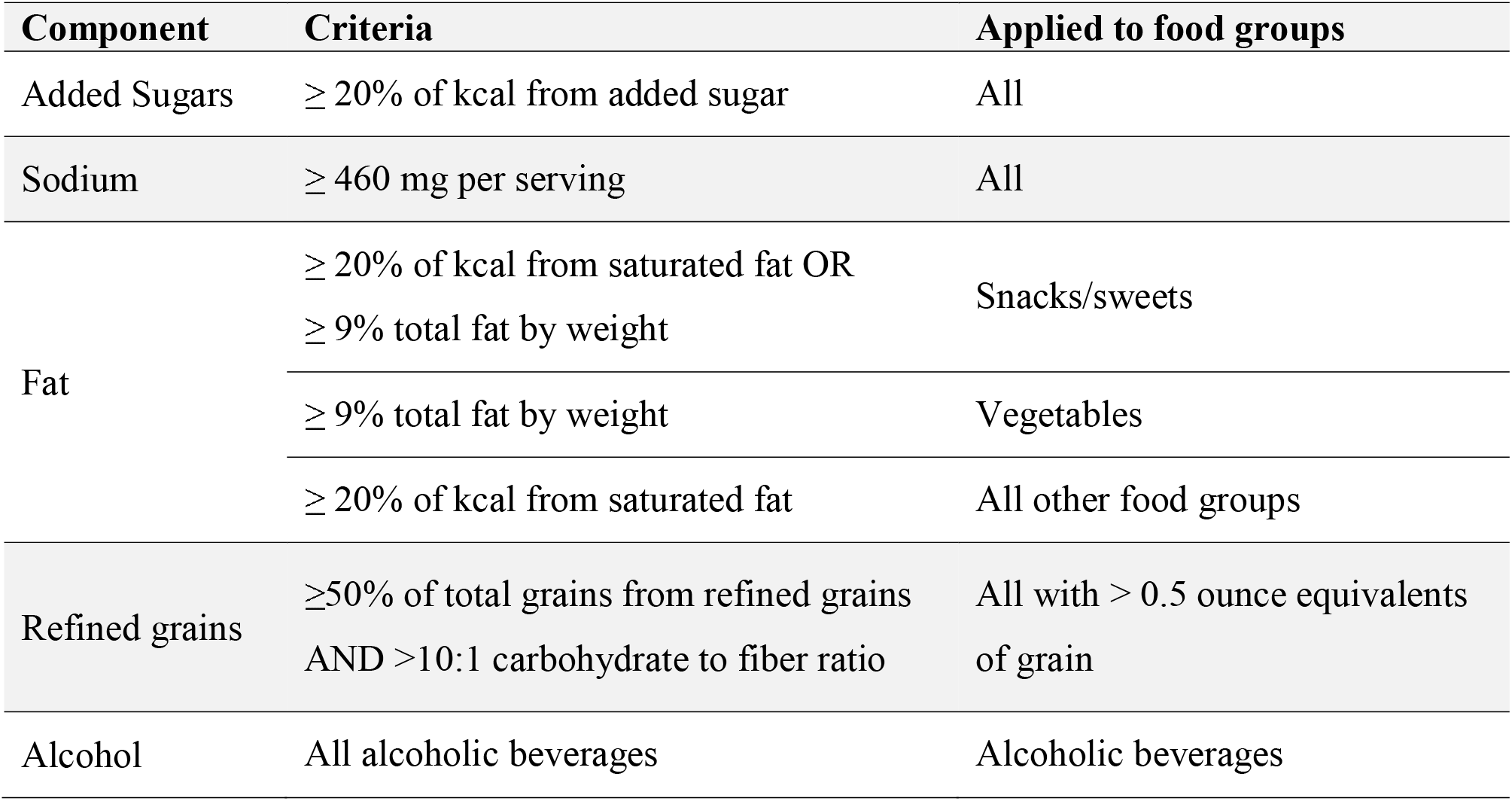
Final criteria for the Moderation Food Classification Method. Foods are classified as moderation if they meet any of the criteria.

Finally, in alignment with the first two guiding principles, it was determined that no whole plant foods (i.e. whole fruit, vegetables, whole grains, legumes, nuts, or seeds) should be classified as a moderation food. ^1^ Only three food items (Brazil nuts, fresh coconut, and avocado [raw or for use on a sandwich]) were misclassified (due to their naturally occurring fat) using the established criteria, so these were recoded manually as non-moderation foods.

### Evaluation of the MFCM

The MFCM was evaluated at both the food and diet levels by assessing its content and construct validity (**Figure 1**). At the food level, content and convergent validity were examined using data from FNDDS (described in more detail below). Content validity was examined by evaluating the accuracy of the method for classifying foods to limit versus foods to encourage as described in the literature, and convergent validity was examined by evaluating the difference in mean dietary components to limit and nutrient density (NRF 9.3 score^56^) of foods classified as moderation versus those classified as non-moderation. At the diet level, internal construct and convergent validity were assessed using WWEIA dietary data (described in more detail below). Internal construct validity was assessed by descriptively examining the proportion of total daily intake of dietary components to limit from moderation foods. Convergent validity was assessed by examining the strength of the relationship of percent of daily energy intake from moderation foods with overall diet quality measured using HEI.

### Data sources

Food- and diet-level validity were assessed using data from the 2017-2018 What We Eat in America (WWEIA), the dietary interview component of the National Health and Nutrition Examination Survey (NHANES), and its linked application data from the Food and Nutrient Database for Dietary Studies. ^29,57^ NHANES is a cross-sectional, nationally representative survey of health and nutrition. Estimates based on weighted samples from each NHANES cycle are representative of the US civilian noninstitutionalized resident population, ^58^ and analyses of recalls are suitable for estimating mean population level intakes from foods and beverages. ^57^ The National Center for Health Statistics Ethics Review Board approves all protocols for NHANES. ^59^ Adult participants provide written consent for their participation; for participants <18 years of age, parents or guardians provide written consent, and documented assent is obtained from participants aged 7-17 years. ^57^

### Food-level nutrient data

The 2017-2018 United States Department of Agriculture (USDA) Food and Nutrient Database for Dietary Studies (FNDDS) ^28^ was used to evaluate the MFCM at the food level. FNDDS is released for each cycle of NHANES and provides nutrient values and gram amounts for foods and beverages reported in the WWEIA dietary intake assessment. The accompanying Food Patterns Equivalents Database (FPED) ^51^ converts the foods and beverages to their corresponding USDA food patterns components (i.e., teaspoon equivalents of added sugars, gram equivalents of saturated fat, and cup equivalents of grains). To facilitate analysis of food and beverage intake, items in these databases are organized into WWEIA food categories based on typical usage and nutrient content. ^60^

The 2017-2018 FNDDS contains nutrient values for 7083 individual foods and beverages consumed in the United States. However, since the moderation food criteria are based on DGA dietary recommendations for individuals over age two years, infant formula and human milk were excluded from these analyses (n=174 foods and beverages). The food weight for “quantity not specified” in the FNDDS Portions and Weights dataset, which reflects the most common portion size^28^ (similar in concept to the FDA reference amount customarily consumed), was used to indicate serving size. For foods with multiple subcodes, serving size was defined as the mean of the portion weights for all subcodes.

### Nationally Representative Dietary Data

WWEIA data was used to evaluate percent energy intake from moderation foods in the US population. WWEIA collects 24-hour dietary recalls from individuals of all ages using the interviewer-administered five-step USDA Automated Multiple-Pass Method. ^57,61^ Dietary recalls are proxy-reported for children 5 years and younger and proxy-assisted for children ages 6-11 years. WWEIA participants also self-report sociodemographic characteristics including age, race, ethnicity, gender, income, and household size. Since the moderation food criteria are based on DGA dietary recommendations for individuals over age two years, the analytic sample for validation included all participants aged two years and older who had two reliable dietary recalls from the 2017-2018 WWEIA cycle. Of 6502 participants with two reliable dietary recalls, 322 participants under two years of age were excluded, resulting in a final sample of 6180 participants. Moderation food intake was calculated as the percentage of total energy intake, pooling across both days.

### Outcome measures

#### Nutrient density

Food-level nutrient density of foods in the 2017-2018 FNDDS^28^ was estimated using the Nutrient Rich Foods Index (NRF) 9.3, which was demonstrated to effectively rank foods by nutrient density and to explain the highest percentage of the variance in the Healthy Eating Index as compared with other NRF formulations. ^56^ The measure is calculated as the sum of the percentages of recommended daily values for nine nutrients to encourage (protein, fiber, vitamin A, vitamin C, Vitamin E, calcium, iron, magnesium, and potassium) minus the percentages of the recommended daily values for three nutrients to limit (saturated fat, added sugars, and sodium), expressed per 100 kcal. Higher scores indicate greater nutrient density. ^56^ ***Diet quality.*** Person-level diet quality was assessed in NHANES participants using the Healthy Eating Index–2020 (HEI), which reflects adherence to the 2020-2025 Dietary Guidelines for Americans. ^1,14^ The HEI total score is calculated as the sum of 13 component scores, including 9 adequacy components (for which higher intake earns higher scores) and 4 moderation components (for which lower intake earns higher scores). HEI^14^ total scores (range 0-100), HEI-adequacy scores (range 0-60), and HEI-moderation scores (range 0-40) were calculated for each NHANES participant using the simple scoring algorithm. ^62^ The HEI-adequacy score is calculated as the sum the component scores for total fruit, whole fruits, total vegetables, greens and beans, whole grains, dairy, total protein foods, seafoods and plant proteins, and fatty acids, while the HEI-moderation score is calculated as the sum of the component scores for refined grains, sodium, added sugars, and saturated fats. Higher total, adequacy, and moderation scores represent closer alignment with the DGA. ^14^ Scores are calculated using density standards (e.g., per 1,000 kilocalories) to account for individual differences in energy requirements. Scores for each component are truncated, whereby maximum scores are assigned when intake meets or exceeds the recommended level for adequacy components, or when intake reaches or falls below a threshold for moderation components; a minimum score of zero is assigned when intake is at or below the minimum threshold for adequacy components, or exceeds the maximum threshold for moderation components.

### Statistical Analysis

The statistical programming code used to implement the MFCM and calculate percent intake from moderation foods is provided at https://github.com/amaracd/Moderation-Food-Classification-Method/. All analyses were completed in SAS 9.4, ^63^ and statistical significance was set at p<0.05. Design-based statistical methods for all NHANES analyses accounted for the complex, multistage sampling design^64^ and were weighted using the dietary recall 2-day sample weights.

### Food-level evaluation

#### Content validity

To operationalize the second guiding principle, content validity at the food level was evaluated descriptively by examining how closely the MFCM food classifications aligned with literature-based characterizations of foods to limit or nutrient-dense foods (**Figure 2**). The percentage of foods classified as moderation foods and the percentage meeting each moderation criterion were summarized overall, by food group, and by WWEIA category in FNDDS. Greater alignment was indicated when most foods in food groups characterized in the literature as those to limit were classified as moderation foods (e.g., sweets, salty snacks, sugar sweetened beverages, high sugar cereal, processed meats, fried potatoes, and high-fat fast food such as pizza, chicken nuggets), ^31–45^ and few foods in food groups referred to as nutrient-dense were classified as moderation foods (e.g., vegetables, fruits, whole grains, seafood, beans, peas, and lentils, unsalted nuts and seeds, eggs, fat-free and low-fat dairy products, and lean meats and poultry that are prepared with minimal amounts of added sugars, saturated fats, and sodium, ^1^). No a priori numerical threshold was specified for acceptable alignment.

#### Convergent validity

Aligned with the first guiding principle and the DGA recommendation to select nutrient-dense versions of foods within each food category, convergent validity was examined at the food level by comparing nutrient density (NRF 9.3 score) of foods classified as moderation versus non-moderation in total and by food group (using t-tests), and across foods meeting 1, 2, and ≥3 MFCM criteria (using ANOVA with Tukey’s HSD post hoc test) in FNDDS (**Figure 1**). In addition, convergent validity was examined by assessing differences in mean energy density and dietary components to limit (i.e. sodium, added sugar, refined grains, carbohydrate to fiber ratio, and saturated fat) between moderation versus non-moderation foods using t-test and across foods meeting 1, 2, and ≥3 MFCM criteria using ANOVA with Tukey’s HSD post hoc test for multiple comparisons. Lower nutrient density, higher energy density, and higher dietary components to limit in moderation versus non-moderation foods indicate strong convergent validity.

### Diet-level evaluation

#### Internal construct validity

Diet-level internal construct validity was evaluated using WWEIA data to describe the proportion of total daily intake of dietary components to limit (i.e. added sugars, saturated fat, refined grains, and sodium). Good internal construct validity was indicated by moderation foods contributing a high proportion of total daily intake of these dietary components.

#### Convergent validity

Convergent validity at the diet level was evaluated by estimating correlations (Pearson’s ρ) of moderation food intake (% kcal) with diet quality (HEI, HEI-moderation, and HEI-adequacy scores). An inverse association of moderation food intake with diet quality indicates convergent validity.

## Results

### Food-level evaluation

The MFCM classified 4555 (66%) of 6909 foods and beverages in the 2017-2018 FNDDS (excluding formula and human milk) as moderation foods (data not shown). Of these, 60% met one criterion, 33% met two criteria, and 7% met three or more criteria. Among foods classified as moderation foods, sodium was the most frequently met MFCM criterion (47%), followed by the refined grain (41%), added sugar (24%), saturated fat (20%), and total fat-by-weight (16%) criteria.

### Food-level content validity

The number and percent of foods classified as moderation foods for each FNDDS food group and WWEIA food category description are presented in **Table 1**.

**Table 1.**
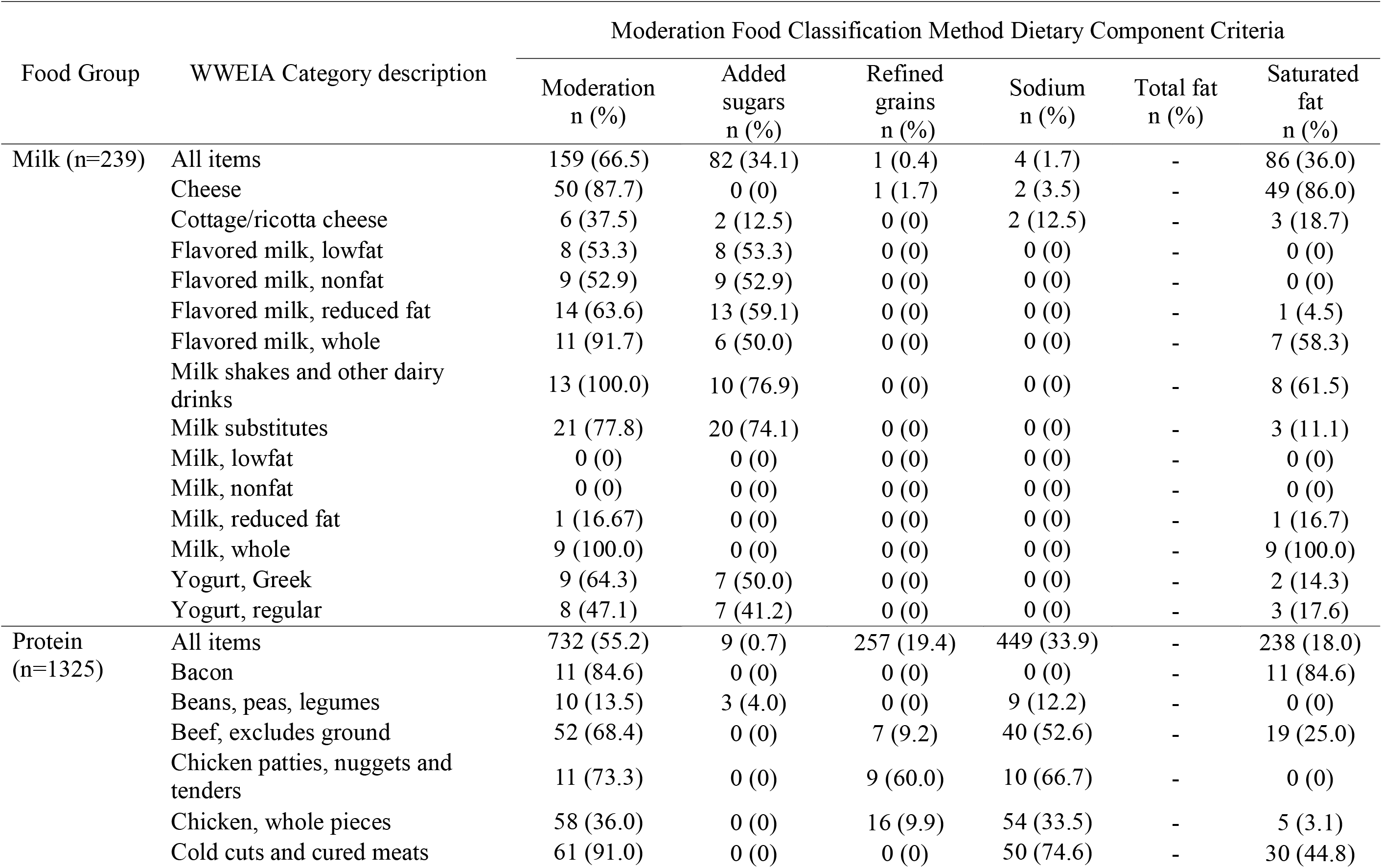
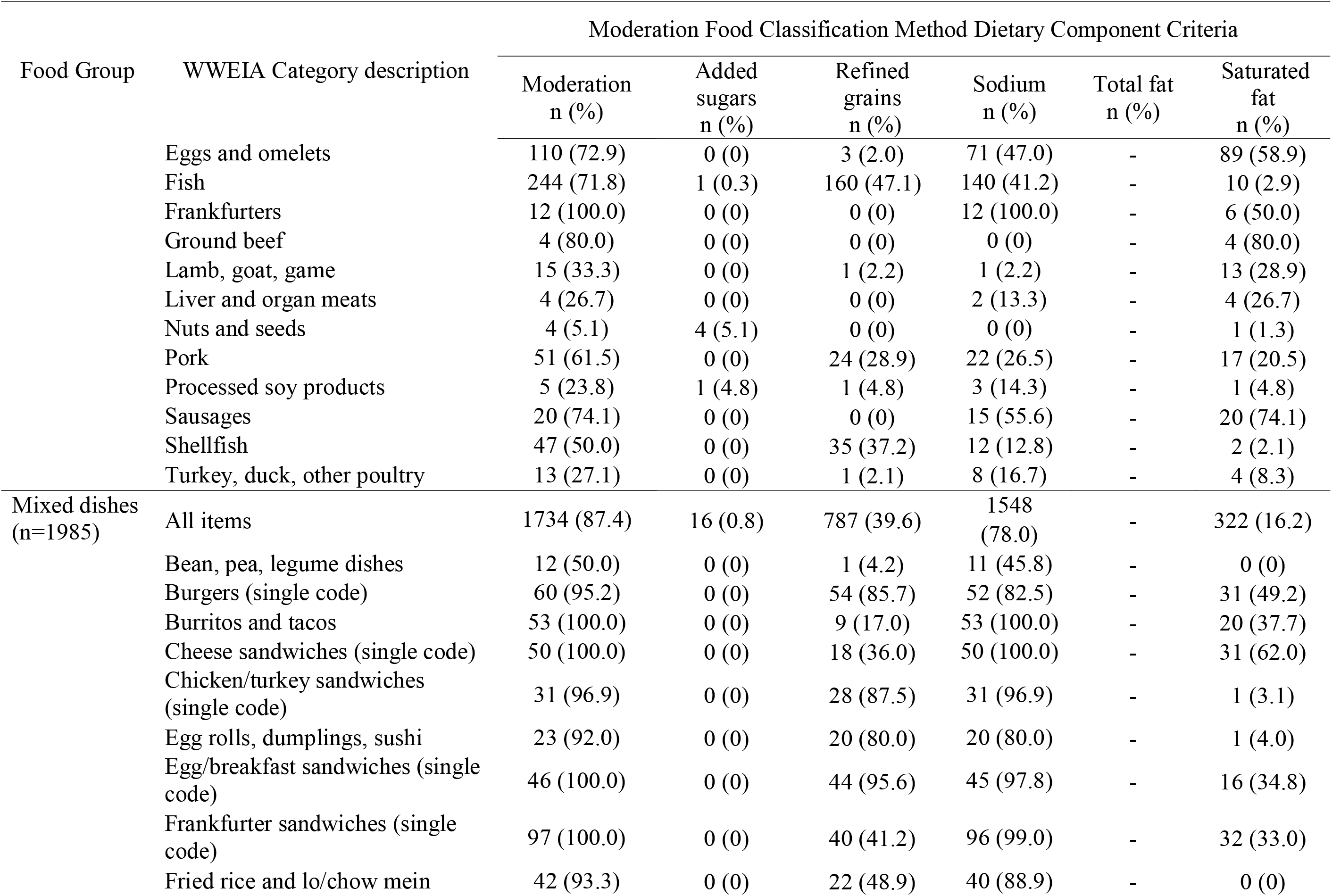
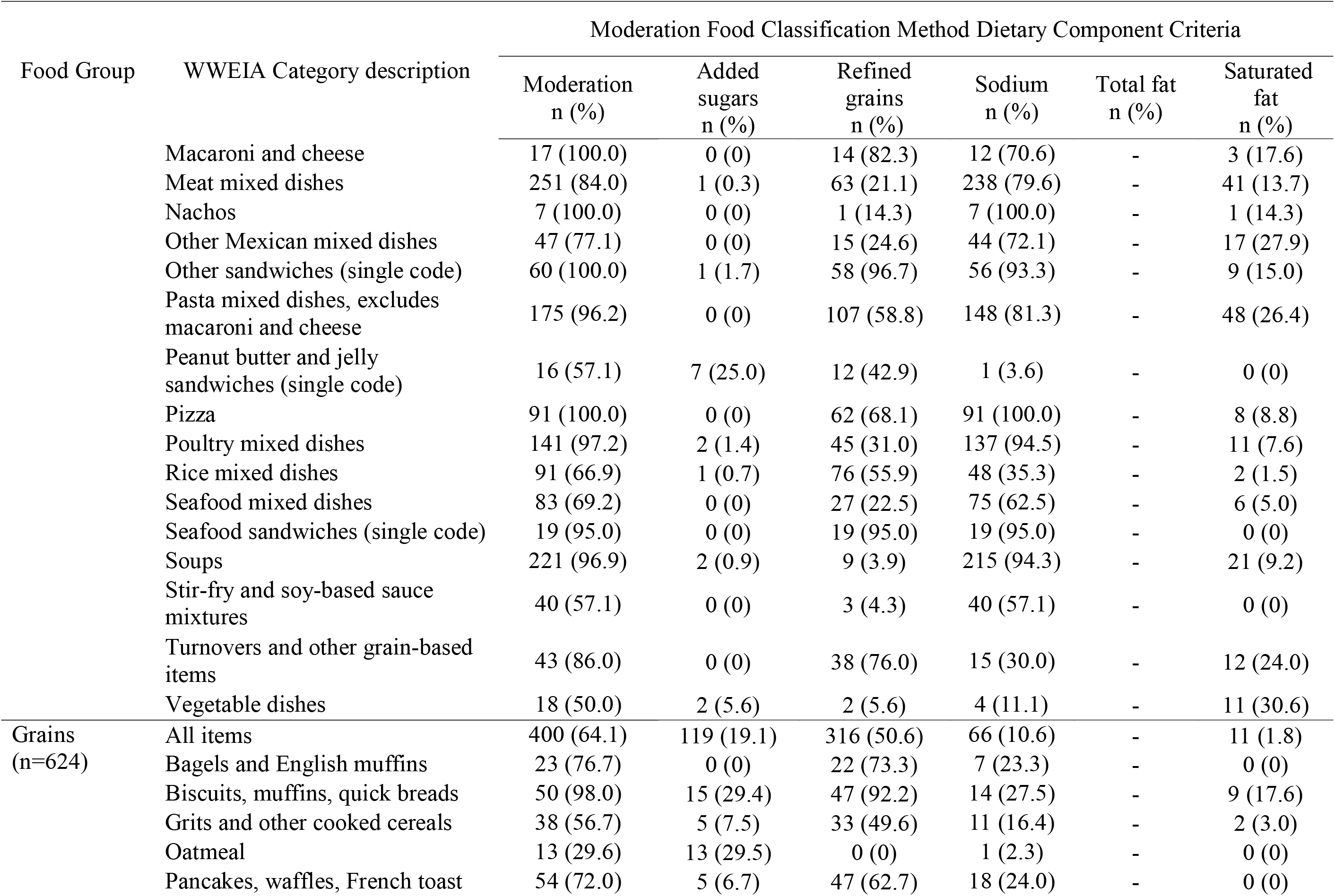
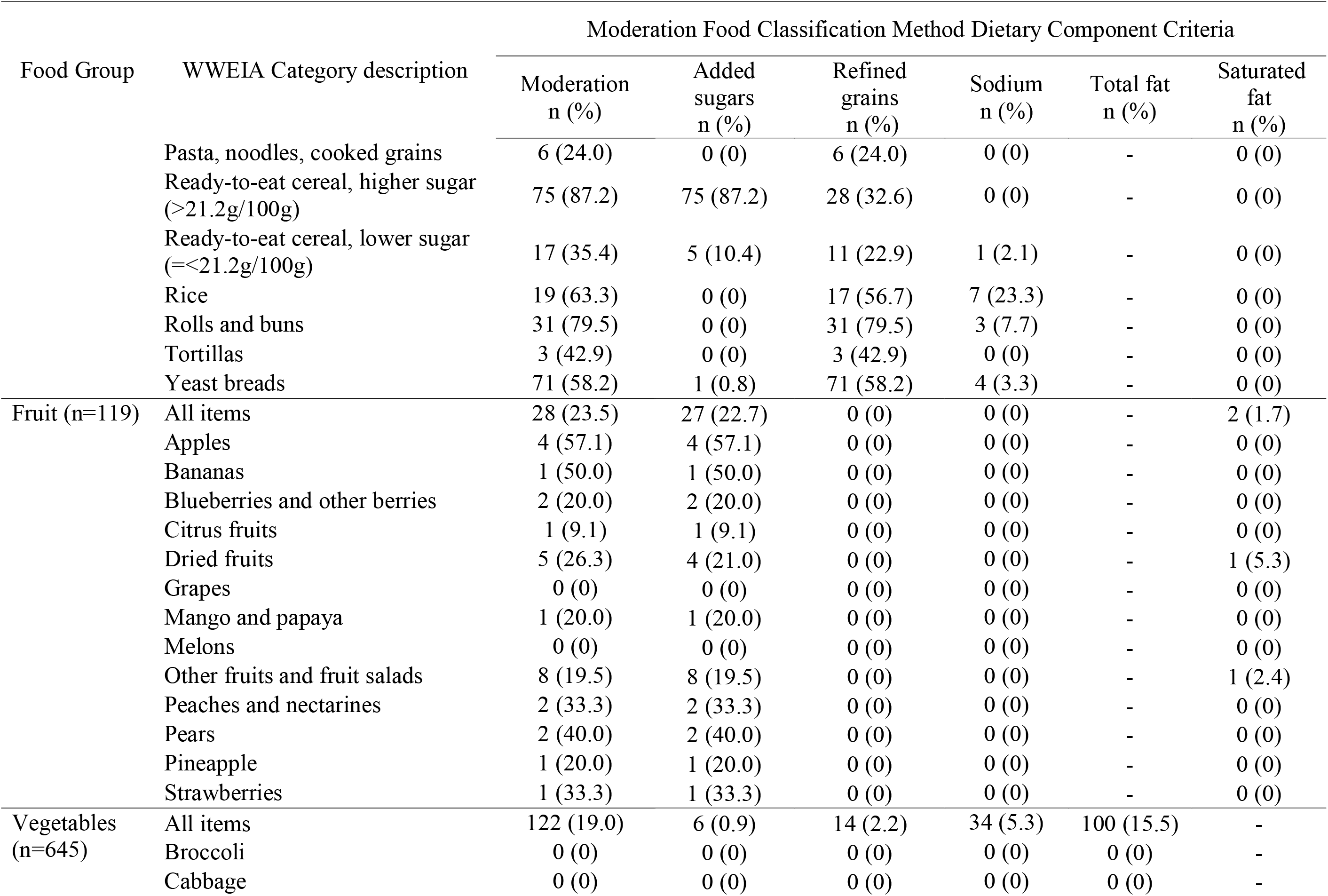
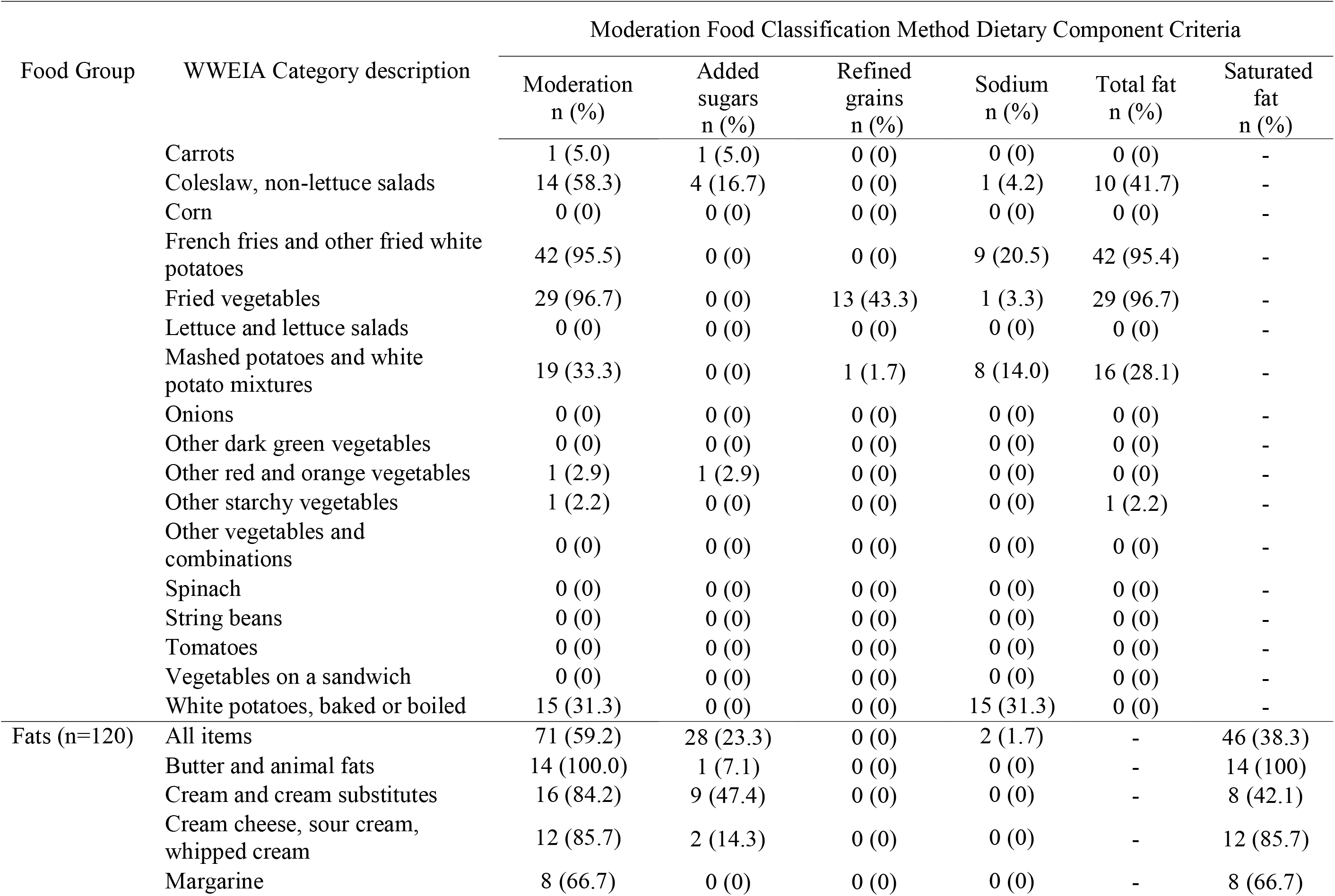
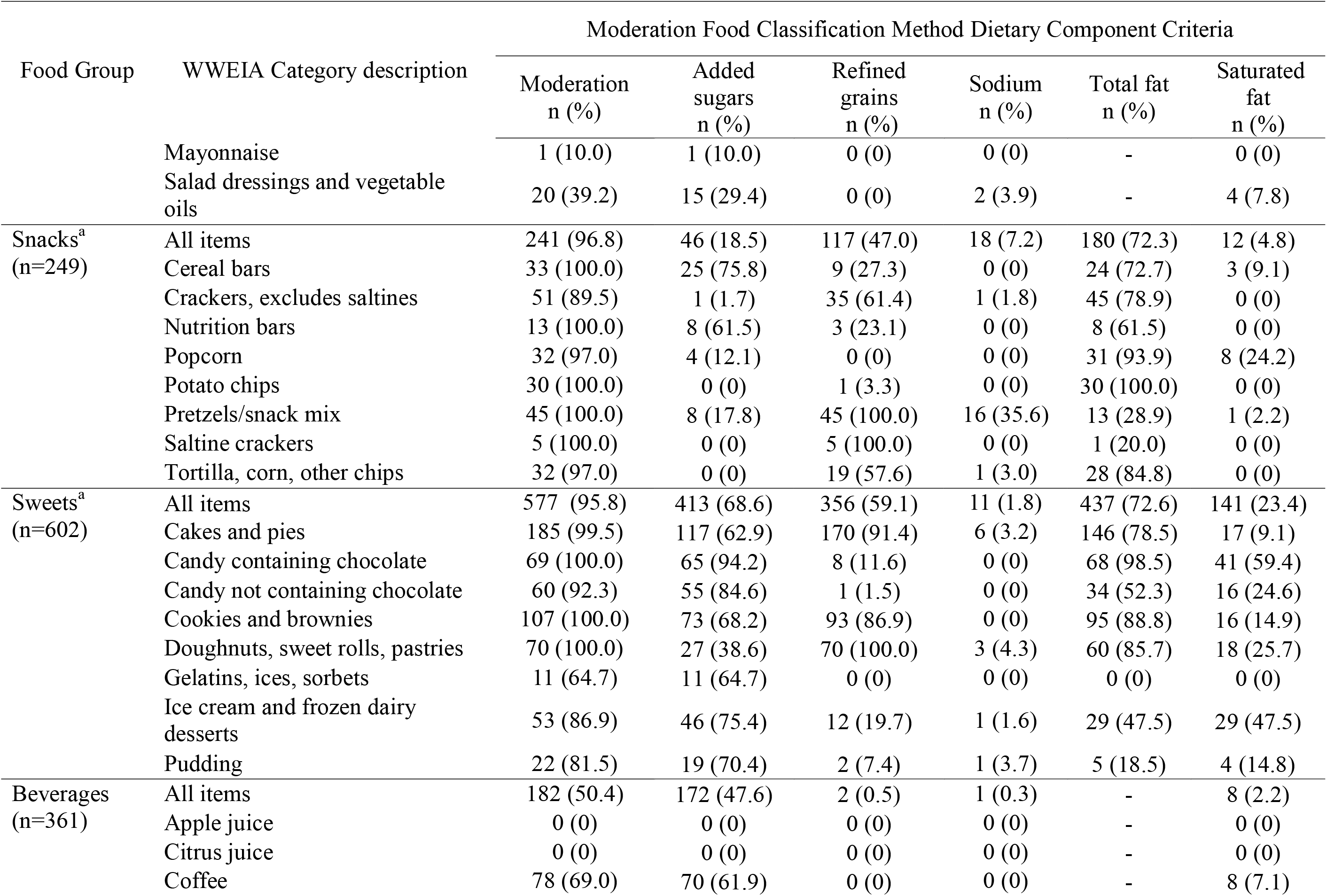
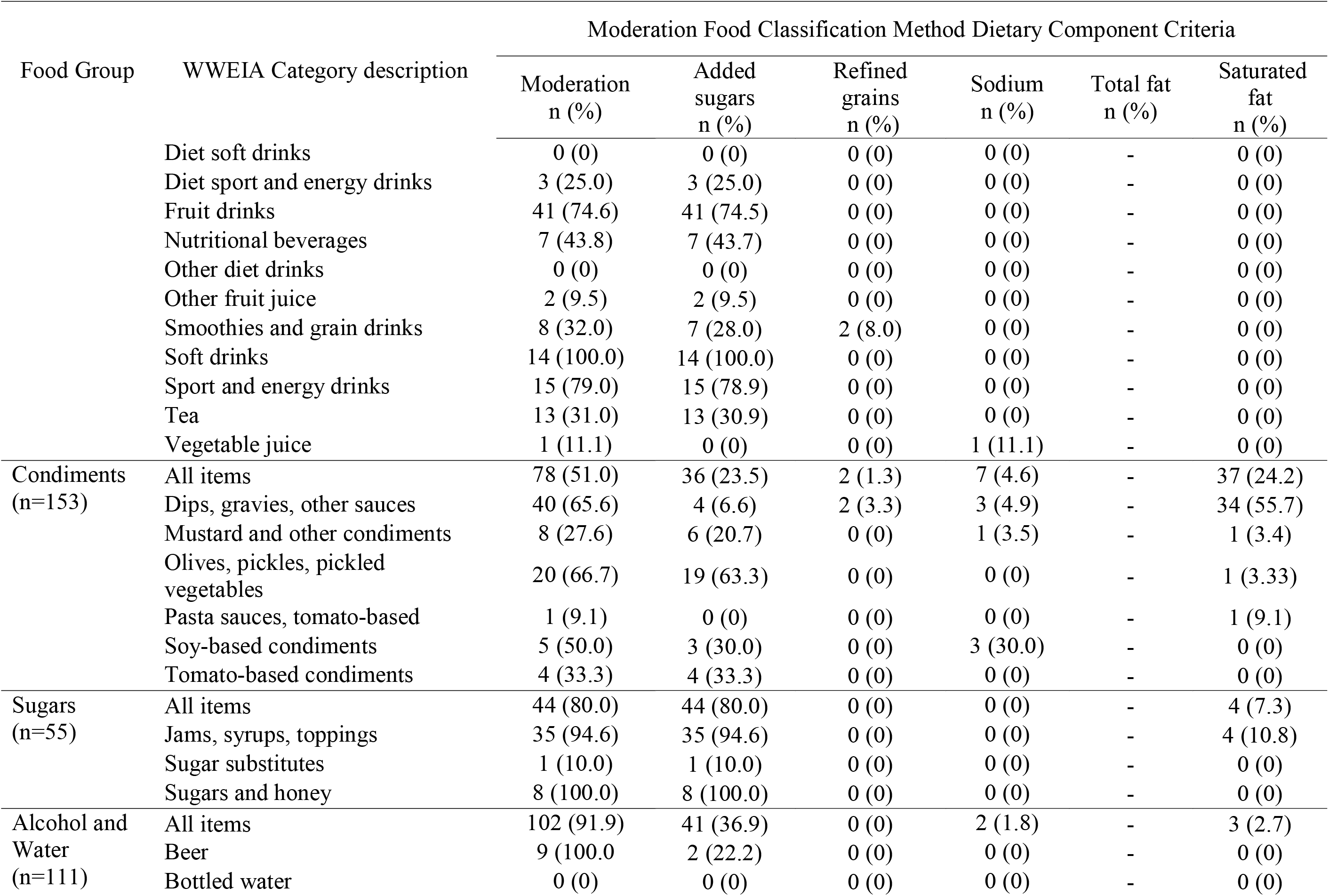
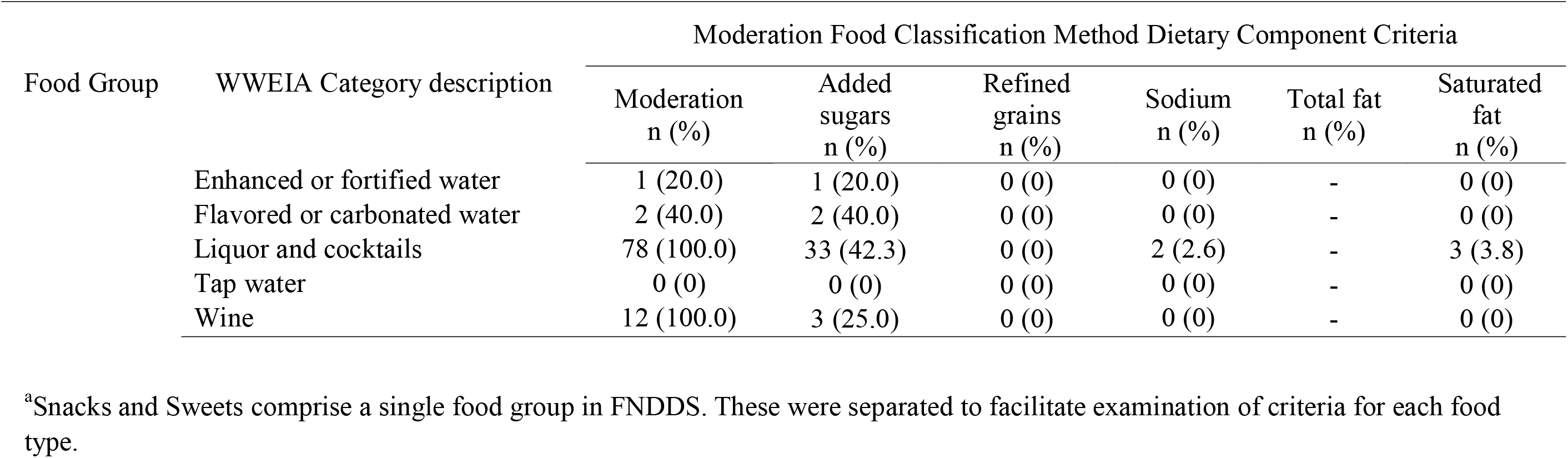
Number and percent of foods classified as moderation and meeting each Moderation Food Classification Method dietary component criterion by What We Eat in America (WWEIA) food category description.

| Food Group | WWEIA Category description | Moderation Food Classification Method Dietary Component Criteria |  |  |  |  |  |
| --- | --- | --- | --- | --- | --- | --- | --- |
|  |  | Moderation<br>n (%) | Added<br>sugars<br>n (%) | Refined<br>grains<br>n (%) | Sodium<br>n (%) | Total fat<br>n (%) | Saturated<br>fat<br>n (%) |
| Milk (n=239) | All items | 159 (66.5) | 82 (34.1) | 1 (0.4) | 4 (1.7) | - | 86 (36.0) |
|  | Cheese | 50 (87.7) | 0 (0) | 1 (1.7) | 2 (3.5) | - | 49 (86.0) |
|  | Cottage/ricotta cheese | 6 (37.5) | 2 (12.5) | 0 (0) | 2 (12.5) | - | 3 (18.7) |
|  | Flavored milk, lowfat | 8 (53.3) | 8 (53.3) | 0 (0) | 0 (0) | - | 0 (0) |
|  | Flavored milk, nonfat | 9 (52.9) | 9 (52.9) | 0 (0) | 0 (0) | - | 0 (0) |
|  | Flavored milk, reduced fat | 14 (63.6) | 13 (59.1) | 0 (0) | 0 (0) | - | 1 (4.5) |
|  | Flavored milk, whole | 11 (91.7) | 6 (50.0) | 0 (0) | 0 (0) | - | 7 (58.3) |
|  | Milk shakes and other dairy<br>drinks | 13 (100.0) | 10 (76.9) | 0 (0) | 0 (0) | - | 8 (61.5) |
|  | Milk substitutes | 21 (77.8) | 20 (74.1) | 0 (0) | 0 (0) | - | 3 (11.1) |
|  | Milk, lowfat | 0 (0) | 0 (0) | 0 (0) | 0 (0) | - | 0 (0) |
|  | Milk, nonfat | 0 (0) | 0 (0) | 0 (0) | 0 (0) | - | 0 (0) |
|  | Milk, reduced fat | 1 (16.67) | 0 (0) | 0 (0) | 0 (0) | - | 1 (16.7) |
|  | Milk, whole | 9 (100.0) | 0 (0) | 0 (0) | 0 (0) | - | 9 (100.0) |
|  | Yogurt, Greek | 9 (64.3) | 7 (50.0) | 0 (0) | 0 (0) | - | 2 (14.3) |
|  | Yogurt, regular | 8 (47.1) | 7 (41.2) | 0 (0) | 0 (0) | - | 3 (17.6) |
| Protein<br>(n=1325) | All items | 732 (55.2) | 9 (0.7) | 257 (19.4) | 449 (33.9) | - | 238 (18.0) |
|  | Bacon | 11 (84.6) | 0 (0) | 0 (0) | 0 (0) | - | 11 (84.6) |
|  | Beans, peas, legumes | 10 (13.5) | 3 (4.0) | 0 (0) | 9 (12.2) | - | 0 (0) |
|  | Beef, excludes ground | 52 (68.4) | 0 (0) | 7 (9.2) | 40 (52.6) | - | 19 (25.0) |
|  | Chicken patties, nuggets and<br>tenders | 11 (73.3) | 0 (0) | 9 (60.0) | 10 (66.7) | - | 0 (0) |
|  | Chicken, whole pieces | 58 (36.0) | 0 (0) | 16 (9.9) | 54 (33.5) | - | 5 (3.1) |
|  | Cold cuts and cured meats | 61 (91.0) | 0 (0) | 0 (0) | 50 (74.6) | - | 30 (44.8) |
|  |  | Moderation<br>n (%) | Added<br>sugars<br>n (%) | Refined<br>grains<br>n (%) | Sodium<br>n (%) | Total fat<br>n (%) | Saturated<br>fat<br>n (%) |
|  | Eggs and omelets | 110 (72.9) | 0 (0) | 3 (2.0) | 71 (47.0) | - | 89 (58.9) |
|  | Fish | 244 (71.8) | 1 (0.3) | 160 (47.1) | 140 (41.2) | - | 10 (2.9) |
|  | Frankfurters | 12 (100.0) | 0 (0) | 0 (0) | 12 (100.0) | - | 6 (50.0) |
|  | Ground beef | 4 (80.0) | 0 (0) | 0 (0) | 0 (0) | - | 4 (80.0) |
|  | Lamb, goat, game | 15 (33.3) | 0 (0) | 1 (2.2) | 1 (2.2) | - | 13 (28.9) |
|  | Liver and organ meats | 4 (26.7) | 0 (0) | 0 (0) | 2 (13.3) | - | 4 (26.7) |
|  | Nuts and seeds | 4 (5.1) | 4 (5.1) | 0 (0) | 0 (0) | - | 1 (1.3) |
|  | Pork | 51 (61.5) | 0 (0) | 24 (28.9) | 22 (26.5) | - | 17 (20.5) |
|  | Processed soy products | 5 (23.8) | 1 (4.8) | 1 (4.8) | 3 (14.3) | - | 1 (4.8) |
|  | Sausages | 20 (74.1) | 0 (0) | 0 (0) | 15 (55.6) | - | 20 (74.1) |
|  | Shellfish | 47 (50.0) | 0 (0) | 35 (37.2) | 12 (12.8) | - | 2 (2.1) |
|  | Turkey, duck, other poultry | 13 (27.1) | 0 (0) | 1 (2.1) | 8 (16.7) | - | 4 (8.3) |
| Mixed dishes<br>(n=1985) | All items | 1734 (87.4) | 16 (0.8) | 787 (39.6) | 1548<br>(78.0) | - | 322 (16.2) |
|  | Bean, pea, legume dishes | 12 (50.0) | 0 (0) | 1 (4.2) | 11 (45.8) | - | 0 (0) |
|  | Burgers (single code) | 60 (95.2) | 0 (0) | 54 (85.7) | 52 (82.5) | - | 31 (49.2) |
|  | Burritos and tacos | 53 (100.0) | 0 (0) | 9 (17.0) | 53 (100.0) | - | 20 (37.7) |
|  | Cheese sandwiches (single code) | 50 (100.0) | 0 (0) | 18 (36.0) | 50 (100.0) | - | 31 (62.0) |
|  | Chicken/turkey sandwiches<br>(single code) | 31 (96.9) | 0 (0) | 28 (87.5) | 31 (96.9) | - | 1 (3.1) |
|  | Egg rolls, dumplings, sushi | 23 (92.0) | 0 (0) | 20 (80.0) | 20 (80.0) | - | 1 (4.0) |
|  | Egg/breakfast sandwiches (single<br>code) | 46 (100.0) | 0 (0) | 44 (95.6) | 45 (97.8) | - | 16 (34.8) |
|  | Frankfurter sandwiches (single<br>code) | 97 (100.0) | 0 (0) | 40 (41.2) | 96 (99.0) | - | 32 (33.0) |
|  | Fried rice and lo/chow mein | 42 (93.3) | 0 (0) | 22 (48.9) | 40 (88.9) | - | 0 (0) |
|  |  | Moderation<br>n (%) | Added<br>sugars<br>n (%) | Refined<br>grains<br>n (%) | Sodium<br>n (%) | Total fat<br>n (%) | Saturated<br>fat<br>n (%) |
|  | Macaroni and cheese | 17 (100.0) | 0 (0) | 14 (82.3) | 12 (70.6) | - | 3 (17.6) |
|  | Meat mixed dishes | 251 (84.0) | 1 (0.3) | 63 (21.1) | 238 (79.6) | - | 41 (13.7) |
|  | Nachos | 7 (100.0) | 0 (0) | 1 (14.3) | 7 (100.0) | - | 1 (14.3) |
|  | Other Mexican mixed dishes | 47 (77.1) | 0 (0) | 15 (24.6) | 44 (72.1) | - | 17 (27.9) |
|  | Other sandwiches (single code) | 60 (100.0) | 1 (1.7) | 58 (96.7) | 56 (93.3) | - | 9 (15.0) |
|  | Pasta mixed dishes, excludes<br>macaroni and cheese | 175 (96.2) | 0 (0) | 107 (58.8) | 148 (81.3) | - | 48 (26.4) |
|  | Peanut butter and jelly<br>sandwiches (single code) | 16 (57.1) | 7 (25.0) | 12 (42.9) | 1 (3.6) | - | 0 (0) |
|  | Pizza | 91 (100.0) | 0 (0) | 62 (68.1) | 91 (100.0) | - | 8 (8.8) |
|  | Poultry mixed dishes | 141 (97.2) | 2 (1.4) | 45 (31.0) | 137 (94.5) | - | 11 (7.6) |
|  | Rice mixed dishes | 91 (66.9) | 1 (0.7) | 76 (55.9) | 48 (35.3) | - | 2 (1.5) |
|  | Seafood mixed dishes | 83 (69.2) | 0 (0) | 27 (22.5) | 75 (62.5) | - | 6 (5.0) |
|  | Seafood sandwiches (single code) | 19 (95.0) | 0 (0) | 19 (95.0) | 19 (95.0) | - | 0 (0) |
|  | Soups | 221 (96.9) | 2 (0.9) | 9 (3.9) | 215 (94.3) | - | 21 (9.2) |
|  | Stir-fry and soy-based sauce<br>mixtures | 40 (57.1) | 0 (0) | 3 (4.3) | 40 (57.1) | - | 0 (0) |
|  | Turnovers and other grain-based<br>items | 43 (86.0) | 0 (0) | 38 (76.0) | 15 (30.0) | - | 12 (24.0) |
|  | Vegetable dishes | 18 (50.0) | 2 (5.6) | 2 (5.6) | 4 (11.1) | - | 11 (30.6) |
| Grains<br>(n=624) | All items | 400 (64.1) | 119 (19.1) | 316 (50.6) | 66 (10.6) | - | 11 (1.8) |
|  | Bagels and English muffins | 23 (76.7) | 0 (0) | 22 (73.3) | 7 (23.3) | - | 0 (0) |
|  | Biscuits, muffins, quick breads | 50 (98.0) | 15 (29.4) | 47 (92.2) | 14 (27.5) | - | 9 (17.6) |
|  | Grits and other cooked cereals | 38 (56.7) | 5 (7.5) | 33 (49.6) | 11 (16.4) | - | 2 (3.0) |
|  | Oatmeal | 13 (29.6) | 13 (29.5) | 0 (0) | 1 (2.3) | - | 0 (0) |
|  | Pancakes, waffles, French toast | 54 (72.0) | 5 (6.7) | 47 (62.7) | 18 (24.0) | - | 0 (0) |
|  |  | Moderation<br>n (%) | Added<br>sugars<br>n (%) | Refined<br>grains<br>n (%) | Sodium<br>n (%) | Total fat<br>n (%) | Saturated<br>fat<br>n (%) |
|  | Pasta, noodles, cooked grains | 6 (24.0) | 0 (0) | 6 (24.0) | 0 (0) | - | 0 (0) |
|  | Ready-to-eat cereal, higher sugar<br>(>21.2g/100g) | 75 (87.2) | 75 (87.2) | 28 (32.6) | 0 (0) | - | 0 (0) |
|  | Ready-to-eat cereal, lower sugar<br>(=<21.2g/100g) | 17 (35.4) | 5 (10.4) | 11 (22.9) | 1 (2.1) | - | 0 (0) |
|  | Rice | 19 (63.3) | 0 (0) | 17 (56.7) | 7 (23.3) | - | 0 (0) |
|  | Rolls and buns | 31 (79.5) | 0 (0) | 31 (79.5) | 3 (7.7) | - | 0 (0) |
|  | Tortillas | 3 (42.9) | 0 (0) | 3 (42.9) | 0 (0) | - | 0 (0) |
|  | Yeast breads | 71 (58.2) | 1 (0.8) | 71 (58.2) | 4 (3.3) | - | 0 (0) |
| Fruit (n=119) | All items | 28 (23.5) | 27 (22.7) | 0 (0) | 0 (0) | - | 2 (1.7) |
|  | Apples | 4 (57.1) | 4 (57.1) | 0 (0) | 0 (0) | - | 0 (0) |
|  | Bananas | 1 (50.0) | 1 (50.0) | 0 (0) | 0 (0) | - | 0 (0) |
|  | Blueberries and other berries | 2 (20.0) | 2 (20.0) | 0 (0) | 0 (0) | - | 0 (0) |
|  | Citrus fruits | 1 (9.1) | 1 (9.1) | 0 (0) | 0 (0) | - | 0 (0) |
|  | Dried fruits | 5 (26.3) | 4 (21.0) | 0 (0) | 0 (0) | - | 1 (5.3) |
|  | Grapes | 0 (0) | 0 (0) | 0 (0) | 0 (0) | - | 0 (0) |
|  | Mango and papaya | 1 (20.0) | 1 (20.0) | 0 (0) | 0 (0) | - | 0 (0) |
|  | Melons | 0 (0) | 0 (0) | 0 (0) | 0 (0) | - | 0 (0) |
|  | Other fruits and fruit salads | 8 (19.5) | 8 (19.5) | 0 (0) | 0 (0) | - | 1 (2.4) |
|  | Peaches and nectarines | 2 (33.3) | 2 (33.3) | 0 (0) | 0 (0) | - | 0 (0) |
|  | Pears | 2 (40.0) | 2 (40.0) | 0 (0) | 0 (0) | - | 0 (0) |
|  | Pineapple | 1 (20.0) | 1 (20.0) | 0 (0) | 0 (0) | - | 0 (0) |
|  | Strawberries | 1 (33.3) | 1 (33.3) | 0 (0) | 0 (0) | - | 0 (0) |
| Vegetables<br>(n=645) | All items | 122 (19.0) | 6 (0.9) | 14 (2.2) | 34 (5.3) | 100 (15.5) | - |
|  | Broccoli | 0 (0) | 0 (0) | 0 (0) | 0 (0) | 0 (0) | - |
|  | Cabbage | 0 (0) | 0 (0) | 0 (0) | 0 (0) | 0 (0) | - |
|  |  | Moderation<br>n (%) | Added<br>sugars<br>n (%) | Refined<br>grains<br>n (%) | Sodium<br>n (%) | Total fat<br>n (%) | Saturated<br>fat<br>n (%) |
|  | Carrots | 1 (5.0) | 1 (5.0) | 0 (0) | 0 (0) | 0 (0) | - |
|  | Coleslaw, non-lettuce salads | 14 (58.3) | 4 (16.7) | 0 (0) | 1 (4.2) | 10 (41.7) | - |
|  | Corn | 0 (0) | 0 (0) | 0 (0) | 0 (0) | 0 (0) | - |
|  | French fries and other fried white potatoes | 42 (95.5) | 0 (0) | 0 (0) | 9 (20.5) | 42 (95.4) | - |
|  | Fried vegetables | 29 (96.7) | 0 (0) | 13 (43.3) | 1 (3.3) | 29 (96.7) | - |
|  | Lettuce and lettuce salads | 0 (0) | 0 (0) | 0 (0) | 0 (0) | 0 (0) | - |
|  | Mashed potatoes and white potato mixtures | 19 (33.3) | 0 (0) | 1 (1.7) | 8 (14.0) | 16 (28.1) | - |
|  | Onions | 0 (0) | 0 (0) | 0 (0) | 0 (0) | 0 (0) | - |
|  | Other dark green vegetables | 0 (0) | 0 (0) | 0 (0) | 0 (0) | 0 (0) | - |
|  | Other red and orange vegetables | 1 (2.9) | 1 (2.9) | 0 (0) | 0 (0) | 0 (0) | - |
|  | Other starchy vegetables | 1 (2.2) | 0 (0) | 0 (0) | 0 (0) | 1 (2.2) | - |
|  | Other vegetables and combinations | 0 (0) | 0 (0) | 0 (0) | 0 (0) | 0 (0) | - |
|  | Spinach | 0 (0) | 0 (0) | 0 (0) | 0 (0) | 0 (0) | - |
|  | String beans | 0 (0) | 0 (0) | 0 (0) | 0 (0) | 0 (0) | - |
|  | Tomatoes | 0 (0) | 0 (0) | 0 (0) | 0 (0) | 0 (0) | - |
|  | Vegetables on a sandwich | 0 (0) | 0 (0) | 0 (0) | 0 (0) | 0 (0) | - |
|  | White potatoes, baked or boiled | 15 (31.3) | 0 (0) | 0 (0) | 15 (31.3) | 0 (0) | - |
| Fats (n=120) | All items | 71 (59.2) | 28 (23.3) | 0 (0) | 2 (1.7) | - | 46 (38.3) |
|  | Butter and animal fats | 14 (100.0) | 1 (7.1) | 0 (0) | 0 (0) | - | 14 (100) |
|  | Cream and cream substitutes | 16 (84.2) | 9 (47.4) | 0 (0) | 0 (0) | - | 8 (42.1) |
|  | Cream cheese, sour cream, whipped cream | 12 (85.7) | 2 (14.3) | 0 (0) | 0 (0) | - | 12 (85.7) |
|  | Margarine | 8 (66.7) | 0 (0) | 0 (0) | 0 (0) | - | 8 (66.7) |
|  |  | Moderation<br>n (%) | Added<br>sugars<br>n (%) | Refined<br>grains<br>n (%) | Sodium<br>n (%) | Total fat<br>n (%) | Saturated<br>fat<br>n (%) |
|  | Mayonnaise | 1 (10.0) | 1 (10.0) | 0 (0) | 0 (0) | - | 0 (0) |
|  | Salad dressings and vegetable oils | 20 (39.2) | 15 (29.4) | 0 (0) | 2 (3.9) | - | 4 (7.8) |
| Snacks<sup>a</sup><br>(n=249) | All items | 241 (96.8) | 46 (18.5) | 117 (47.0) | 18 (7.2) | 180 (72.3) | 12 (4.8) |
|  | Cereal bars | 33 (100.0) | 25 (75.8) | 9 (27.3) | 0 (0) | 24 (72.7) | 3 (9.1) |
|  | Crackers, excludes saltines | 51 (89.5) | 1 (1.7) | 35 (61.4) | 1 (1.8) | 45 (78.9) | 0 (0) |
|  | Nutrition bars | 13 (100.0) | 8 (61.5) | 3 (23.1) | 0 (0) | 8 (61.5) | 0 (0) |
|  | Popcorn | 32 (97.0) | 4 (12.1) | 0 (0) | 0 (0) | 31 (93.9) | 8 (24.2) |
|  | Potato chips | 30 (100.0) | 0 (0) | 1 (3.3) | 0 (0) | 30 (100.0) | 0 (0) |
|  | Pretzels/snack mix | 45 (100.0) | 8 (17.8) | 45 (100.0) | 16 (35.6) | 13 (28.9) | 1 (2.2) |
|  | Saltine crackers | 5 (100.0) | 0 (0) | 5 (100.0) | 0 (0) | 1 (20.0) | 0 (0) |
|  | Tortilla, corn, other chips | 32 (97.0) | 0 (0) | 19 (57.6) | 1 (3.0) | 28 (84.8) | 0 (0) |
| Sweets<sup>a</sup><br>(n=602) | All items | 577 (95.8) | 413 (68.6) | 356 (59.1) | 11 (1.8) | 437 (72.6) | 141 (23.4) |
|  | Cakes and pies | 185 (99.5) | 117 (62.9) | 170 (91.4) | 6 (3.2) | 146 (78.5) | 17 (9.1) |
|  | Candy containing chocolate | 69 (100.0) | 65 (94.2) | 8 (11.6) | 0 (0) | 68 (98.5) | 41 (59.4) |
|  | Candy not containing chocolate | 60 (92.3) | 55 (84.6) | 1 (1.5) | 0 (0) | 34 (52.3) | 16 (24.6) |
|  | Cookies and brownies | 107 (100.0) | 73 (68.2) | 93 (86.9) | 0 (0) | 95 (88.8) | 16 (14.9) |
|  | Doughnuts, sweet rolls, pastries | 70 (100.0) | 27 (38.6) | 70 (100.0) | 3 (4.3) | 60 (85.7) | 18 (25.7) |
|  | Gelatins, ices, sorbets | 11 (64.7) | 11 (64.7) | 0 (0) | 0 (0) | 0 (0) | 0 (0) |
|  | Ice cream and frozen dairy desserts | 53 (86.9) | 46 (75.4) | 12 (19.7) | 1 (1.6) | 29 (47.5) | 29 (47.5) |
|  | Pudding | 22 (81.5) | 19 (70.4) | 2 (7.4) | 1 (3.7) | 5 (18.5) | 4 (14.8) |
| Beverages<br>(n=361) | All items | 182 (50.4) | 172 (47.6) | 2 (0.5) | 1 (0.3) | - | 8 (2.2) |
|  | Apple juice | 0 (0) | 0 (0) | 0 (0) | 0 (0) | - | 0 (0) |
|  | Citrus juice | 0 (0) | 0 (0) | 0 (0) | 0 (0) | - | 0 (0) |
|  | Coffee | 78 (69.0) | 70 (61.9) | 0 (0) | 0 (0) | - | 8 (7.1) |

|  |  | Moderation Food Classification Method Dietary Component Criteria |  |  |  |  |  |
| --- | --- | --- | --- | --- | --- | --- | --- |
| Food Group | WWEIA Category description | Moderation<br>n (%) | Added<br>sugars<br>n (%) | Refined<br>grains<br>n (%) | Sodium<br>n (%) | Total fat<br>n (%) | Saturated<br>fat<br>n (%) |
|  | Diet soft drinks | 0 (0) | 0 (0) | 0 (0) | 0 (0) | - | 0 (0) |
|  | Diet sport and energy drinks | 3 (25.0) | 3 (25.0) | 0 (0) | 0 (0) | - | 0 (0) |
|  | Fruit drinks | 41 (74.6) | 41 (74.5) | 0 (0) | 0 (0) | - | 0 (0) |
|  | Nutritional beverages | 7 (43.8) | 7 (43.7) | 0 (0) | 0 (0) | - | 0 (0) |
|  | Other diet drinks | 0 (0) | 0 (0) | 0 (0) | 0 (0) | - | 0 (0) |
|  | Other fruit juice | 2 (9.5) | 2 (9.5) | 0 (0) | 0 (0) | - | 0 (0) |
|  | Smoothies and grain drinks | 8 (32.0) | 7 (28.0) | 2 (8.0) | 0 (0) | - | 0 (0) |
|  | Soft drinks | 14 (100.0) | 14 (100.0) | 0 (0) | 0 (0) | - | 0 (0) |
|  | Sport and energy drinks | 15 (79.0) | 15 (78.9) | 0 (0) | 0 (0) | - | 0 (0) |
|  | Tea | 13 (31.0) | 13 (30.9) | 0 (0) | 0 (0) | - | 0 (0) |
|  | Vegetable juice | 1 (11.1) | 0 (0) | 0 (0) | 1 (11.1) | - | 0 (0) |
| Condiments<br>(n=153) | All items | 78 (51.0) | 36 (23.5) | 2 (1.3) | 7 (4.6) | - | 37 (24.2) |
|  | Dips, gravies, other sauces | 40 (65.6) | 4 (6.6) | 2 (3.3) | 3 (4.9) | - | 34 (55.7) |
|  | Mustard and other condiments | 8 (27.6) | 6 (20.7) | 0 (0) | 1 (3.5) | - | 1 (3.4) |
|  | Olives, pickles, pickled<br>vegetables | 20 (66.7) | 19 (63.3) | 0 (0) | 0 (0) | - | 1 (3.33) |
|  | Pasta sauces, tomato-based | 1 (9.1) | 0 (0) | 0 (0) | 0 (0) | - | 1 (9.1) |
|  | Soy-based condiments | 5 (50.0) | 3 (30.0) | 0 (0) | 3 (30.0) | - | 0 (0) |
|  | Tomato-based condiments | 4 (33.3) | 4 (33.3) | 0 (0) | 0 (0) | - | 0 (0) |
| Sugars<br>(n=55) | All items | 44 (80.0) | 44 (80.0) | 0 (0) | 0 (0) | - | 4 (7.3) |
|  | Jams, syrups, toppings | 35 (94.6) | 35 (94.6) | 0 (0) | 0 (0) | - | 4 (10.8) |
|  | Sugar substitutes | 1 (10.0) | 1 (10.0) | 0 (0) | 0 (0) | - | 0 (0) |
|  | Sugars and honey | 8 (100.0) | 8 (100.0) | 0 (0) | 0 (0) | - | 0 (0) |
| Alcohol and<br>Water<br>(n=111) | All items | 102 (91.9) | 41 (36.9) | 0 (0) | 2 (1.8) | - | 3 (2.7) |
|  | Beer | 9 (100.0) | 2 (22.2) | 0 (0) | 0 (0) | - | 0 (0) |
|  | Bottled water | 0 (0) | 0 (0) | 0 (0) | 0 (0) | - | 0 (0) |
|  |  | Moderation<br>n (%) | Added<br>sugars<br>n (%) | Refined<br>grains<br>n (%) | Sodium<br>n (%) | Total fat<br>n (%) | Saturated<br>fat<br>n (%) |
|  | Enhanced or fortified water | 1 (20.0) | 1 (20.0) | 0 (0) | 0 (0) | - | 0 (0) |
|  | Flavored or carbonated water | 2 (40.0) | 2 (40.0) | 0 (0) | 0 (0) | - | 0 (0) |
|  | Liquor and cocktails | 78 (100.0) | 33 (42.3) | 0 (0) | 2 (2.6) | - | 3 (3.8) |
|  | Tap water | 0 (0) | 0 (0) | 0 (0) | 0 (0) | - | 0 (0) |
|  | Wine | 12 (100.0) | 3 (25.0) | 0 (0) | 0 (0) | - | 0 (0) |

<sup>a</sup>Snacks and Sweets comprise a single food group in FNDDS. These were separated to facilitate examination of criteria for each food type.

#### Classification within food groups to limit

The MCFM classified greater than 95% of snacks and sweets as moderation foods. Within the WWEIA categories, all milkshakes, frankfurters, pizza, nachos, potato chips, cookies and brownies, doughnuts, and soft drinks were classified as moderation foods. Additionally, at least 85% of the processed meats (i.e. bacon, cold cuts), high sugar cereals, and fried potatoes were classified as moderation foods. The sweets group contained the highest number of foods that met two component criteria (50%; data not shown). Only snacks (4%), mixed dishes (6%), and sweets (28%) contained more than 2% of foods that met three or more criteria.

#### Classification within food groups to encourage

The MCFM classified 19% of foods in the vegetable group and 24% of foods in the fruit group as moderation foods. In the vegetable group, most foods classified as moderation met the total fat criteria. In the fruit group, most foods classified as moderation met the criteria for added sugar (**Table 1)**. Within the WWEIA categories, 13% of beans, peas, and lentils, 5% of nuts and seeds, and none of the low-fat or non-fat milk, grapes, melon, broccoli, spinach, string beans, or tomatoes were classified as moderation foods. The vegetable categories that contained more than 30% moderation foods included coleslaw, mashed potatoes, white potatoes, and fried potatoes or vegetables. In the protein food group, 36% of foods in the chicken category and 27% of turkey, duck, and other poultry were classified as moderation foods, primarily due to exceeding criteria for sodium.

### Food-level convergent validity

Mean energy density, sodium, carbohydrate to fiber ratio, percent grains from refined grain, and percent kcal from added sugar and saturated fat were higher in moderation foods compared to non-moderation foods (p<0.001; **Table 2**). Mean energy density, carbohydrate to fiber ratio, and percent kcal from saturated fat increased with the number of moderation criteria met (p<0.05; **Table 2**). Mean percent kcal from added sugar was higher for foods meeting one and three criteria than for those meeting two criteria (p<0.05). Mean sodium and percent grains from refined grains increased with the number of moderation criteria met, although there was no significant difference between those meeting two versus three criteria.

**Table 2.**
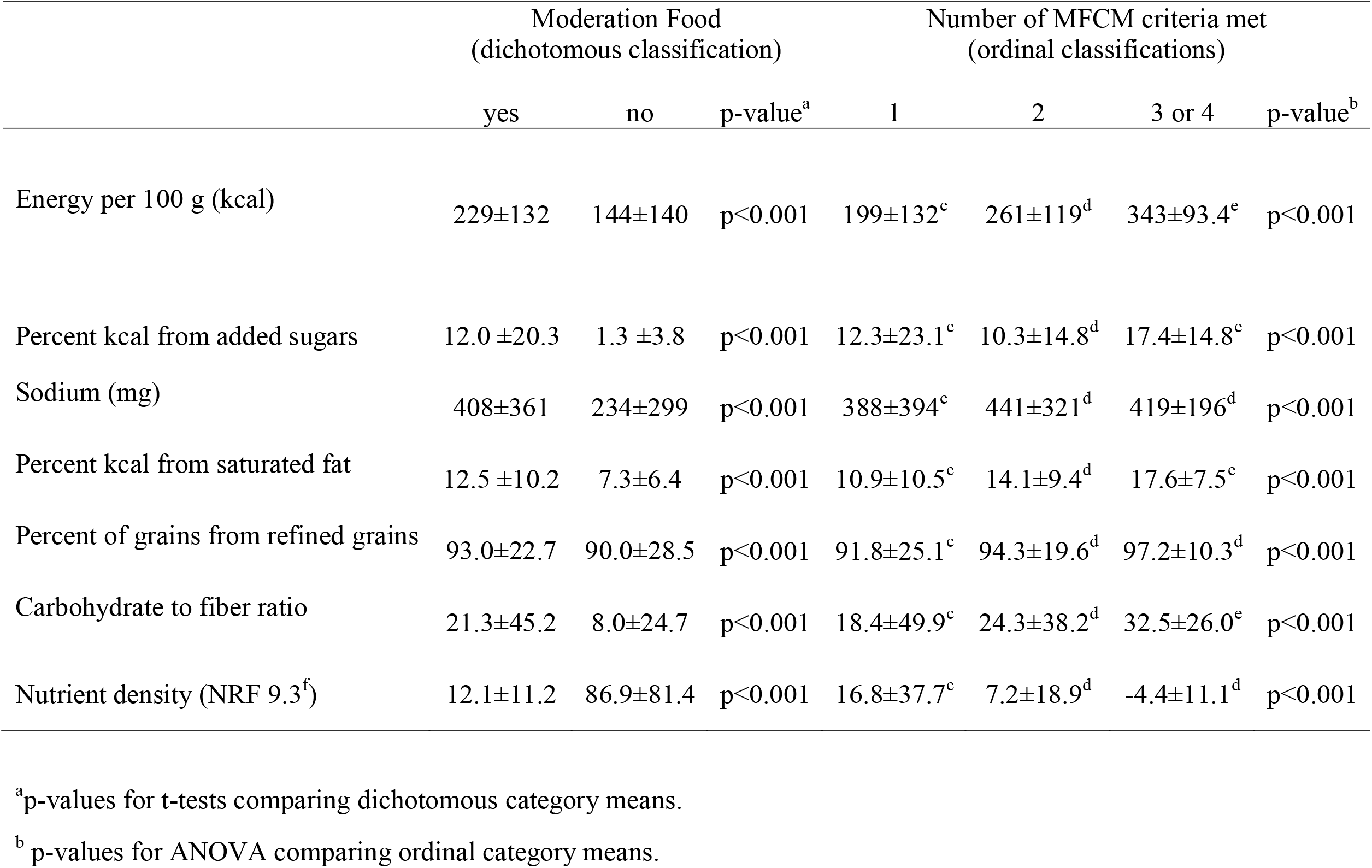
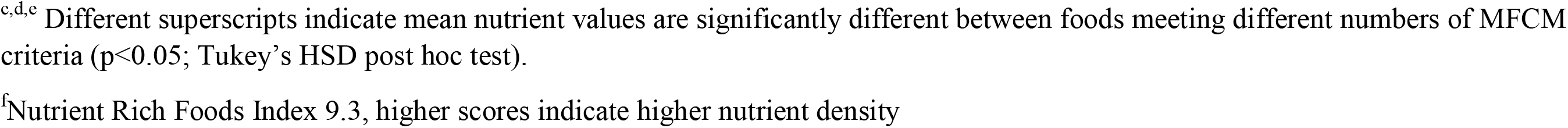
Summary of dietary components to limit and nutrient density (mean±SD) by dichotomous and ordinal categories based on the Moderation Food Classification Method (MFCM) in the 2017-2018 Food and Nutrient Database for Dietary Studies (FNDDS; n=6909).

|  | Moderation Food<br>(dichotomous classification) |  |  | Number of MFCM criteria met<br>(ordinal classifications) |  |  |  |
| --- | --- | --- | --- | --- | --- | --- | --- |
|  | yes | no | p-value<sup>a</sup> | 1 | 2 | 3 or 4 | p-value<sup>b</sup> |
| Energy per 100 g (kcal) | 229±132 | 144±140 | p<0.001 | 199±132<sup>c</sup> | 261±119<sup>d</sup> | 343±93.4<sup>e</sup> | p<0.001 |
| Percent kcal from added sugars | 12.0 ±20.3 | 1.3 ±3.8 | p<0.001 | 12.3±23.1<sup>c</sup> | 10.3±14.8<sup>d</sup> | 17.4±14.8<sup>e</sup> | p<0.001 |
| Sodium (mg) | 408±361 | 234±299 | p<0.001 | 388±394<sup>c</sup> | 441±321<sup>d</sup> | 419±196<sup>d</sup> | p<0.001 |
| Percent kcal from saturated fat | 12.5 ±10.2 | 7.3±6.4 | p<0.001 | 10.9±10.5<sup>c</sup> | 14.1±9.4<sup>d</sup> | 17.6±7.5<sup>e</sup> | p<0.001 |
| Percent of grains from refined grains | 93.0±22.7 | 90.0±28.5 | p<0.001 | 91.8±25.1<sup>c</sup> | 94.3±19.6<sup>d</sup> | 97.2±10.3<sup>d</sup> | p<0.001 |
| Carbohydrate to fiber ratio | 21.3±45.2 | 8.0±24.7 | p<0.001 | 18.4±49.9<sup>c</sup> | 24.3±38.2<sup>d</sup> | 32.5±26.0<sup>e</sup> | p<0.001 |
| Nutrient density (NRF 9.3<sup>f</sup>) | 12.1±11.2 | 86.9±81.4 | p<0.001 | 16.8±37.7<sup>c</sup> | 7.2±18.9<sup>d</sup> | -4.4±11.1<sup>d</sup> | p<0.001 |

<sup>a</sup>p-values for t-tests comparing dichotomous category means. <sup>b</sup> p-values for ANOVA comparing ordinal category means.
<sup>c,d,e</sup> Different superscripts indicate mean nutrient values are significantly different between foods meeting different numbers of MFCM criteria ( $p < 0.05$ ; Tukey's HSD post hoc test). <sup>f</sup> Nutrient Rich Foods Index 9.3, higher scores indicate higher nutrient density

Boxplots were used to visualize differences in nutrient density (i.e. NRF 9.3 scores) of moderation vs. non-moderation foods overall (**Figure 5A**), by number of criteria met (**Figure 5B**), and for each of the individual food groups (**Supplemental Figure 6**). Mean nutrient density was significantly lower in moderation foods than non-moderation foods overall (p<0.001; **Table 2**). Mean nutrient density decreased progressively with increasing number of moderation criteria met, although there was no significant difference between those meeting two versus three criteria (p<0.001; **Table 2**).

**Figure 5.**
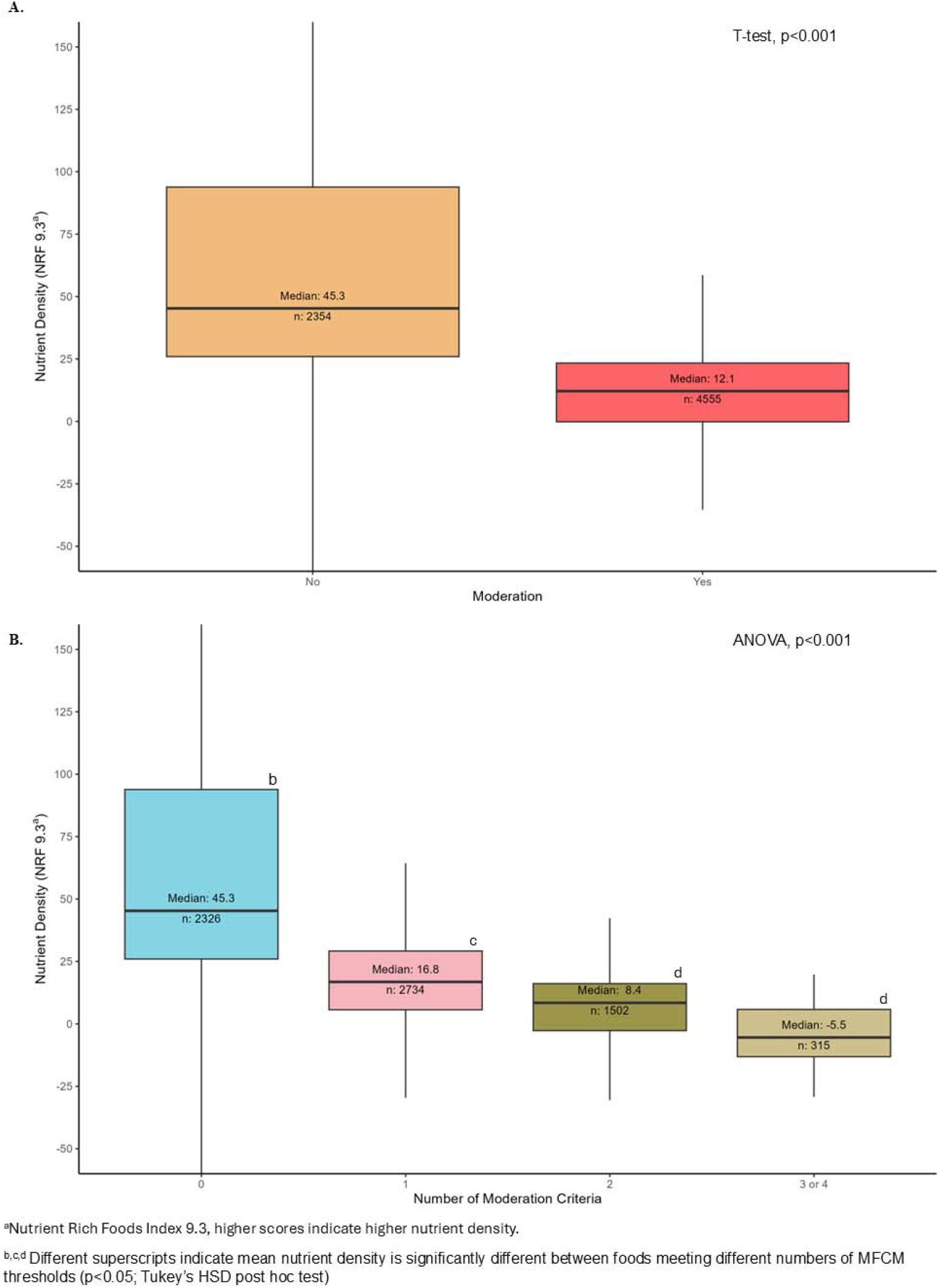
**A**) Nutrient density for moderation versus non-moderation foods according to the Moderation Food Classification Method. **B**) Nutrient density according to number of moderation thresholds met.

### Diet-level evaluation

The 2017-2018 NHANES sample was majority non-Hispanic White and 51% female. The mean age was 39 years (SE: 0.5) and mean IPR was 2.9 (SE:0.1) (**Table 3**).

**Table 3.** Survey-Weighted Analytic Sample Characteristics (n=6180) and Dietary Intake Estimates in the 2017-2018 What We Eat in America (WWEIA)

| **Participant characteristic** | **Mean ± SE or n (%)<sup>a</sup>** |
| --- | --- |
| Age (years) | 39.4 ± 0.5 |
| Race and Ethnicity |  |
| Hispanic Mexican American | 869 (11) |
| Hispanic other | 512 (7) |
| Non-Hispanic Asian | 700 (5) |
| Non-Hispanic Black | 1514 (12) |
| Non-Hispanic White | 2170 (59) |
| Other race - including multi-racial | 415 (5) |
| Gender |  |
| Male | 2974 (49) |
| Female | 3206 (51) |
| Family Income to Poverty Ratio<sup>b</sup> | 2.9 ± 0.1 |
| **Dietary intake estimate** | **Mean ± SE<sup>a</sup>** |
| Moderation food intake (% kcal) | 76.9 ± 0.6 |
| Intake from foods meeting 1 MFCM criterion (% kcal) | 42.0±0.3 |
| Intake from foods meeting 2 MFCM criteria (% kcal) | 26.2±0.4 |
| Intake from foods meeting 3 or 4 MFCM criteria (% kcal) | 8.7±0.3 |
| % teaspoon equivalents of added sugar intake attributable to moderation foods | 94.3±0.4 |
| % mg of sodium intake attributable to moderation foods | 78.3±0.5 |
| % ounce equivalents of refined grain intake attributable to moderation foods | 94.4±0.3 |
| % grams of Saturated fat intake attributable to moderation foods | 82.6±0.5 |

<sup>a</sup>Means and weighted percentages were estimated accounting for the complex survey design.
<sup>b</sup>Family income to federal poverty threshold, with higher values reflecting higher income.

### Diet-level internal construct validity

Weighted mean proportion of intake attributable to moderation foods was 77% (SE:0.6) of total daily energy intake (**Table 3**). Foods exceeding only one moderation food criterion contributed 42% of total daily energy intake (SE: 0.3), while foods exceeding two criteria contributed 26% of total daily energy intake (SE:0.4), and foods exceeding three or more criteria contributed 9% of total daily energy intake (SE: 0.3). Moderation foods contributed 94% of total daily added sugar intake (SE: 0.4), 78% of total daily sodium intake (SE:0.5), 83% of total daily saturated fat (SE: 0.5), and 94% of total daily refined grains (SE:0.3).

### Diet-level convergent validity

Intake from moderation foods was negatively correlated with HEI total, HEI-moderation, and HEI-adequacy scores (**Table 4**).

**Table 4.** Pearson correlation of moderation food intake (% kcal) with Healthy Eating Index-2020 (HEI) scores in participants 2 years of age and older (n=6180) in the 2017-2018 What We Eat in America (WWEIA).

|  | 1 | 2 | 3 | 4 |
| --- | --- | --- | --- | --- |
| 1. Moderation food intake (% kcal) | - |  |  |  |
| 2. HEI<sup>a</sup> | -0.72*** | - |  |  |
| 3. HEI Moderation<sup>b</sup> | -0.56*** | 0.81*** | - |  |
| 4. HEI Adequacy<sup>c</sup> | -0.67*** | 0.92*** | 0.51*** | - |

\*\*\*p<0.0001
<sup>a</sup>Healthy Eating Index-2020 total score, higher scores indicate greater adherence to 2020-2025 Dietary Guidelines for Americans.
<sup>b</sup>Healthy Eating Index moderation score, calculated as the sum of the 4 moderation components, higher scores indicate lower intake.
<sup>c</sup>Healthy Eating Index adequacy score, calculated as the sum of the 9 adequacy components, higher scores indicate higher intake.

## Discussion

This study developed a quantitative method applying standardized criteria to classify moderation foods, i.e., foods that should be consumed in moderation due to containing high amounts of dietary components to limit, aligned with the DGA. ^1^ The MFCM demonstrated high food-level content validity, as evidenced by the large proportion of moderation foods in the snacks (97%), sweets (96%), and mixed dishes (87%) food groups, and the small proportion of moderation foods in the vegetable (19%) and fruit food groups (23%), consistent with the second guiding principle. The MFCM demonstrated strong convergent validity at the food level, as indicated by the significantly lower nutrient density in moderation versus non-moderation foods (overall and within each food group) and the inverse relationship of the number of moderation criteria met with nutrient density. The measure demonstrated strong diet-level internal construct validity, as evidenced by the high proportion of daily intake from target dietary components attributed to moderation foods. The MFCM also demonstrated strong convergent validity at the person level, as evidenced by the negative association of moderation food intake (% kcal) with HEI scores.

Food classifications from the MFCM criteria closely align with foods to limit described in prior literature, ^31–45,65^ and included the top food contributors to added sugar, ^66,67^ saturated fat, ^67,68^ and sodium^69^ in the U.S. diet. Of note, snack foods were more often classified as moderation foods based on the fat and refined grain criteria than the sodium criterion. This likely reflects their relatively small portion sizes, such that the sodium content per portion did not exceed the criterion. However, this is consistent with national intake data showing that mixed and main dishes contribute more dietary sodium than snacks, the latter accounting for approximately 4% of total sodium intake. ^69^

Although the positive association of moderation food intake with the HEI-moderation components score indicates alignment with the DGA, the magnitude of the association (r = −0.56) suggests non-overlapping variance between the two scores. Moderation food intake calculated using this food-based method may more precisely reflect intake of nutrient-poor foods than the HEI-moderation score, ^14^ as it does not truncate intake and it accounts for some dietary components not explicitly captured in the HEI-moderation score, such as unsaturated fat from chips and many sweets. Additionally, while the HEI-moderation score includes dietary components to limit that come from core foods (e.g., saturated fat from low-fat milk or lean meat^70^), these are not counted as moderation food intake using the food-based method. Future research elucidating whether moderation food intake and HEI-moderation scores are differentially related to health outcomes would be informative.

The MFCM was designed to classify foods that are major contributors to the excess intake of dietary components of concern in the U.S. population and therefore should be consumed in moderation. Classifying foods as moderation versus not may be useful for guiding consumer behavior, product labeling, and quantifying the contribution of moderation food intake to overall dietary intake when examining diet-disease relationships. However, any binary food classification will include foods of varying nutritional value within each classification. The inverse association of nutrient density with the number of criteria met indicates there is variance in the nutritional value of moderation foods. Similarly, foods not classified as moderation may also vary in their nutritional value and contribution to DGA guidelines, and should be consumed within daily energy requirements across all recommended food groups.

The MFCM has utility for research, practice, and policy. This evidence-based, quantitative method can be applied across widely varying diets and allows for comparison of findings across studies of diverse populations. The ability to classify individual moderation foods may also facilitate clinical dietary education, ^13^ actionable consumer guidance, and product labeling. For example, in 2016, the Chilean government mandated warning labels on foods using nutrient thresholds similar to the current moderation food classification. The policy led to decreased intakes of sugar, sodium, and saturated fat, ^71,72^ suggesting that clearly labeled foods can improve eating behaviors. In addition, the MFCM could be used to monitor and evaluate consumer purchasing behaviors and the availability of moderation foods in the food supply, such as in restaurants or schools.

### Strengths and limitations

Strengths of this study include the use of evidence-based criteria applied to the FNDDS database, ^28^ which supports the internal validity and generalizability of the classification method. The use of FNDDS for classifying moderation foods is a strength, as it is a comprehensive database for foods consumed by the U.S. population and enables implementation of the classification method with data from NHANES WWEIA^29^ or the National Cancer Institute Automated Self-Administered 24-hour dietary assessment tool (ASA24®).^30^ However, as the criteria were developed for FNDSS, they will likely require adaptation for use in studies based on other nutrient databases. Another strength is the use of a large, nationally representative sample of the civilian, noninstitutionalized U.S. population to estimate associations of moderation food intake with overall diet quality. Limitations include the use of self-reported dietary intake data, which are susceptible to response bias. However, this is not expected to impact comparisons of moderation food intake with HEI scores, as misreporting similarly impact the independent and dependent variables. Food-level analyses are also limited to the foods included in FNDDS, in which items represent generic food representations instead of specific brands. Consequently, differences in recipe and preparation methods that alter nutrient content may not be fully captured. However, these issues would similarly impact any classification-based content of dietary components. Additionally, there is variability in how the MFCM criteria were established because of the difficulty of directly applying HEI guidance to individual foods. Nevertheless, the criteria consistently identify lower nutrient-dense foods and demonstrate strong convergent validity.

### Conclusions

These findings support the validity of the MFCM for identifying foods with lower nutrient density and for quantifying moderation food intake, which is inversely related to adherence to dietary guidelines in the U.S. population. By establishing quantitative dietary-component-based criteria that effectively identify foods high in dietary components to limit, the MFCM has utility for diet quality assessment, behavioral nutrition interventions, and investigations of diet-disease relationships.

## Data Availability

All data produced in the present study are available upon reasonable request.

---

**Supplemental Figure 6.**
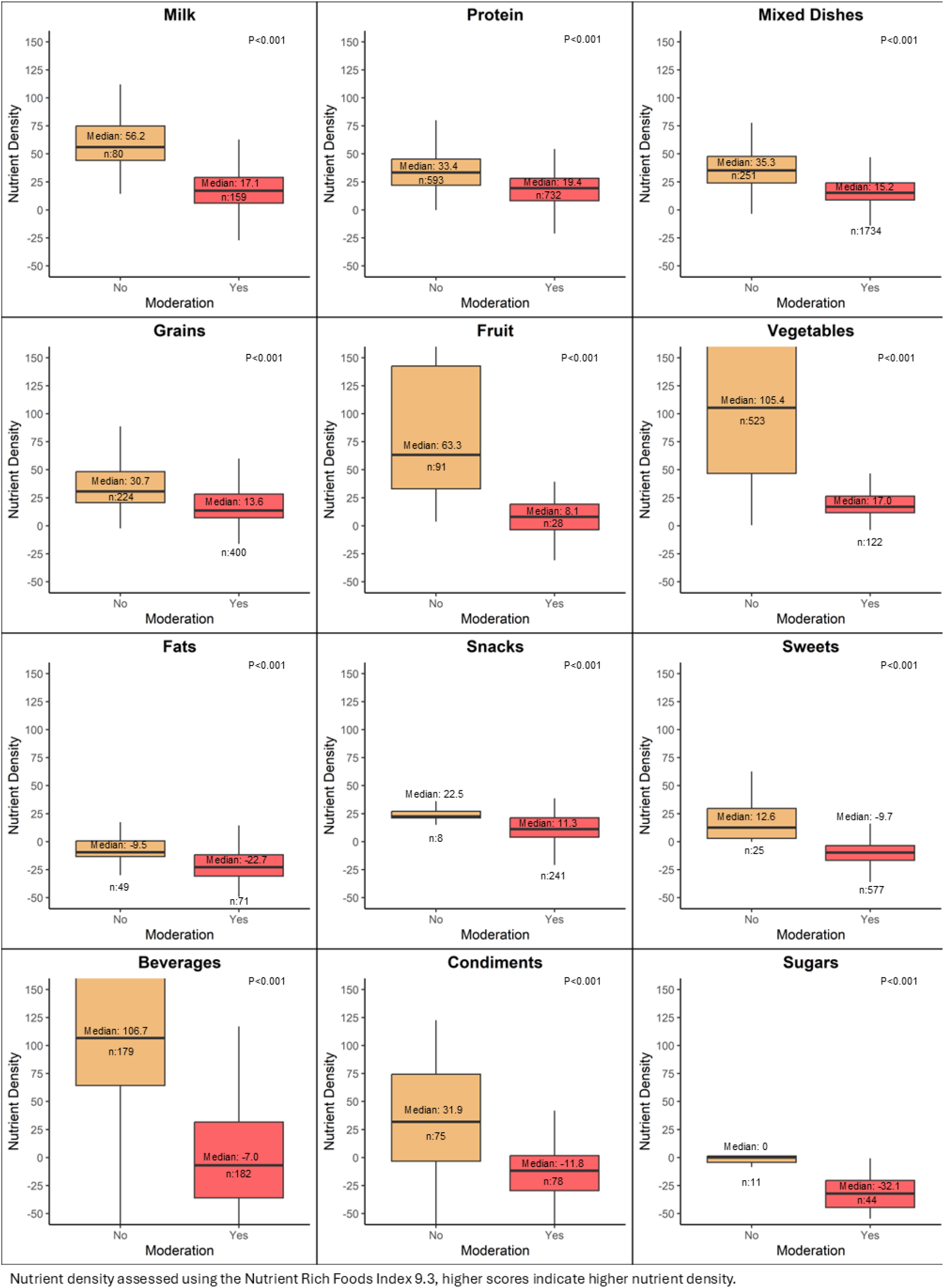
Nutrient density by moderation food classification for each food group.

